# Reference values and validation of the 1-min sit-to-stand test in healthy 5- to 16-year-old youth: a cross-sectional study

**DOI:** 10.1101/2021.01.14.21249789

**Authors:** Sarah R Haile, Thea Fühner, Urs Granacher, Julien Stocker, Thomas Radtke, Susi Kriemler

**Author notes:** shared first authorship (Sarah R Haile, Thea Fühner). shared last authorship (Thomas Radtke, Susi Kriemler). **Corresponding author and contact details** Prof. Dr. Susi Kriemler, Epidemiology, Biostatistics and Prevention Institute, University of Zurich, Hirschengraben 84, 8001 Zurich, Switzerland.

## Abstract

**Objectives:** It is essential to have simple, reliable and valid tests to measure children’s functional capacity in schools or medical practice. The 1-min sit-to-stand test (STS) is a quick fitness test requiring little equipment or space that is increasingly used in both healthy populations and those with chronic disease. We aimed to provide age and sex-specific reference values of STS in healthy children and adolescents and to evaluate its short-term reliability and construct validity.

**Design, setting and participants:** Cross-sectional random sample from 6 public schools and 1 science fair in central Europe. Overall, 587 healthy participants aged 5-16 years were recruited and divided into age groups of 3 years each.

**Outcomes:** 1-minute STS. To evaluate short-term reliability, some children performed the STS twice. To evaluate construct validity, some children also performed a standing long jump (SLJ) and a maximal incremental exercise test.

**Results:** Data from 547 5-16 year old youth were finally included in the analyses. The median number of repetitions in 1 minute in males (females) ranged from 55 [95% CI 38 to 72] (53 [35 to 76]) in 14-16 year-olds to 59 [41 to 77] (60 [38 to 77]) in 8-10 year-olds. Children who repeated STS showed a learning effect of on average 4.8 repetitions more than the first test (95% limits of agreement −6.7 to 16.4). Moderate correlations were observed between the STS and the SLJ (*r* = 0.48) and the maximal exercise test (*r* = 0.43).

**Conclusions:** The reported STS reference values can be used to interpret STS test performance in children and adolescents. The STS appears to have good test-retest reliability, but a learning effect of about 10%. The association of STS with other measures of physical fitness should be further explored in a larger study and technical standards for its conduct are needed.

**Strengths and Limitations of this Study:**

- Large sample size (N = 547)
- Reference values according to sex and age group (5-7, 8-10, 11-13 and 14-16)
- Evaluation of test-retest reliability and construct validity
- Convenience not population-based sample
- Not all outcomes have been measured on each participant

## Introduction

Physical fitness is widely understood to be an indicator of an individual’s overall health status as well as an important predictor of morbidity and mortality [1, 2]. A wide range of fitness tests have been developed for the assessment of muscular strength (e.g., grip strength test, standing long jump test [SLJ], and isokinetic dynamometry [3–6]) and aerobic capacity (e.g., 6-min walk test [6MWT], cardiopulmonary exercise on a treadmill or stationary bike [CPET], and 20-meter multi-stage fitness test [7–9]) in healthy individuals as well as in patients suffering from chronic diseases [2].

One increasingly used functional capacity test is the 1-min sit-to-stand test (STS) [10], which evaluates how many times per minute an individual is able to stand up and sit down on a chair standardized for height [2]. The STS is an attractive alternative to other tests for the assessment of overall physical fitness as it is simple, quick, requires only a chair and a stopwatch, and is possible even in small spaces, indicating that it can equally be used as an exercise test or a screening instrument. It has previously been evaluated in adults, especially those with chronic conditions such as chronic obstructive pulmonary disease (COPD) [1, 11] or cystic fibrosis [7]. Increasing interest has been shown to the use of STS as a test of functional exercise capacity in children, both healthy [12, 13] and with chronic conditions [14]. In small samples, STS has been observed to correlate well with the 6MWT [14] and with final heart rate [13] during the test.

In recent years, several studies provided reference values for the STS in adults, but to our knowledge no age-specific standard values are available for healthy children and adolescents [2]. Standard values are needed to interpret the exercise capacity of children and youth, both healthy and with various diseases, in a wide range of settings. Some prediction models and overall references including large age ranges, however, exist [13]. Data concerning construct validity and test-retest reliability in healthy children and adolescents are also lacking.

We aimed to measure STS performance in healthy children and adolescents in order to provide age and sex specific reference values, to assess short-term reliability, and to evaluate construct validity. Based on the adult literature [15], we hypothesized that the STS is reliable and valid in youth and that performance increases with age and lean mass. To examine the construct validity of STS, we compared repetitions of STS with other commonly used functional capacity outcomes including standing long jump and a bicycle test.

## Methods

### Study Design

This cross-sectional study was comprised of a convenience sample of children and adolescents aged 5-16 years. For the evaluation of short-term reliability, children and adolescents performed the STS twice, with a break of 15 minutes between trials. To test construct validity, a subsample of children and adolescents additionally performed a maximal incremental exercise test on a cycle ergometer as a measure of aerobic capacity and the SLJ as a measure of muscular strength.

### Population

A convenience sample of healthy children and adolescents aged 5-16 years from five public schools located in three different Swiss towns (Biel, Grenchen and Zurich) were contacted in January 2017 and asked to participate. In addition, one public school located in Potsdam, Germany participated in September 2019. Furthermore, we recruited children and adolescents during Scientifica, an open-door Swiss science fair organized by the University of Zurich and the Swiss Federal Institute of Technology in Zurich (ETH Zurich) in September 2017. We aimed to enroll at least 70 children per age category (5-7, 8-10, 11-13, 14-16 years), based on feasibility considerations.

Test results were stored anonymously in a database of the Epidemiology, Biostatistics and Prevention Institute (EBPI) of the University of Zurich, Switzerland. The Ethical Committee of the Canton of Zurich waived the need for ethical approval, as this study does not fall under the scope of the Human Research Act in Switzerland [16]. Written informed consent (i.e., signature by participant or parent/caregiver) is not mandatory since the study measurements included only minimal risks for the study participants and the data were collected anonymously. Swiss children did however provide their oral consent to participate. In Germany, all participants and their legal guardians were informed about potential risks and benefits of the study prior to enrollment and legal guardians provided their written informed consent. The protocol was approved by the local ethical commission of the University of Potsdam (submission No. 45/2019).

### Measurements

In order to estimate age and sex-specific reference values, all children performed at least one STS. For the evaluation of short-term reliability, some participants performed a second STS. To test construct validity, a subset of children also performed either the maximal incremental exercise test for the assessment of aerobic capacity or the SLJ for the assessment of muscle strength.

#### Assessment of anthropometrics

Standing height (in cm, accuracy 0.5 cm), leg length (from the anterior inferior iliac spine to the lateral malleolus in 5 cm, accuracy 1 cm), and body mass (in kg, accuracy 100 g) were assessed. Body mass index (BMI) was calculated and z-scores derived [17]. During the assessment of anthropometrics, children and adolescents wore sportswear and gym shoes (sneakers).

#### 1-min sit-to-stand test

In the STS, children and adolescents had to stand up and sit down on a chair without arm rests as many times as possible within one minute (working instructions in Supplementary Material 1). The wooden chair was adapted to the individual height of the children, so that the angle of their knee joints was approximately 90° while sitting. Furthermore, the chair height (in cm) from the sitting plane to the floor was measured. During the test, participants had to keep their feet parallel and to put their hands on their waist in order to ensure that they would not use their arms to assist the movement [2, 8]. They were instructed to straighten their knees and stand up completely, and to touch the chair with the buttocks when sitting down. The instructor told the participants to complete as many sit-to-stand repetitions as possible within one minute. They were informed when 15 seconds were left, but they were not motivated by the instructor during the test. The number of fully completed and correct STS cycles after one minute was recorded for the analysis. The STS at schools was conducted by a single member of the study team. At Scientifica, six different people conducted the STS precisely adhering to standard operational procedures.

#### Maximal incremental exercise test on a stationary bike

A subset of children and adolescents completed an exercise test on a children’s bicycle ergometer (Corival Pediatric, Lode, NL). The exercise test was based on the Godfrey protocol [18] in which the children started at a different workload depending on their standing height and the workload was increased every one minute depending on their standing height (i.e., height <120 cm =10 W/min, height 120-150 cm = 15 W/min, and > 150 cm = 20 W/min) until exhaustion. During the test, the children wore a heart rate monitor (Polar RCX5TM). The children were told to pedal at ≥ 60 revolutions per minute (rpm). The test was stopped as soon as at least one of the following criteria was reached: subjective exhaustion ≥ 8 on a 0-10 modified Borg scale [19], heart rate ≥ 190 beats/minute, or the participants could not maintain a pedaling speed of at least 60 rpm. Maximal workload (Wmax) and maximal workload adjusted for body mass (Wmax / kg) were computed for the analysis.

#### Standing long jump

In the SLJ [6], the children had to jump as far as they could in horizontal direction from an upright standing position. An arm swing prior to and during the jump was allowed. The jump distance between the starting line and the heel of the posterior foot was retained for the analysis. SLJ was repeated three times, of which the furthest distance was recorded as best trial. To adjust for children’s differing heights, SLJ performance was analyzed as best distance divided by height.

### Statistical Analysis

Anthropometric data and STS data were summarized as n and median [interquartile range] according to sex and age group (5-7, 8-10, 11-13, 14-16 years) and reported using percentiles 2.5%, 25%, 50%, 75%, 97.5%. We considered 4 definitions of STS outcome: best of 2 tests, first STS, last STS, and mean first and second STS. Test-retest reliability between first and second STS was assessed using the method of Bland and Altman [20], where mean difference in the two tests assesses their agreement, along with 95% limits of agreement. Construct validity was examined by comparing STS to SLJ and maximal exercise test using Pearson’s correlation coefficient. We also considered partial correlations of the three anthropometric measures (i.e., standing height, leg length, and body mass) adjusting for age. Correlation coefficients of 0-0.19 were considered very weak, 0.2-0.39 weak, 0.4-0.59 moderate, 0.60-0.79 strong and 0.80–1.0 as very strong [21]. The statistical analysis was performed using the R programming language [22] (R Version 4.0.3), and code is available in Supplementary Material 2.

## Results

### Study Population

Of the 587 participants, we excluded 13 because they were younger than 5 or older than 16, 8 had missing age or STS, and a further 19 were unmotivated to perform the tests, leaving a final number of 547 subjects. We divided the study population into four different age groups: 5-7 years, 8-10 years, 11-13 years and 14-16 years. Descriptive characteristics of each age group are shown in Table 1. Overall, 373 (68%) participants performed the STS twice, 43 (8%) performed the maximal exercise test, and 72 (13%) did the SLJ (Supplementary Table S1).

**Table 1.** Study population including all participants aged 5-16 years with at least one sit-to-stand test. Values are median and [interquartile range].

| Characteristic | 5-7 years<br>n = 122 (♀ 59) | 8-10 years<br>n = 160 (♀ 88) | 11-13 years<br>n = 165 (♀ 75) | 14-16 years<br>n = 100 (♀ 47) | all<br>n = 547 (♀ 269) |
| --- | --- | --- | --- | --- | --- |
| Standing height [m] | 1.26 [1.22, 1.30] | 1.39 [1.34, 1.45] | 1.57 [1.52, 1.64] | 1.67 [1.63, 1.74] | 1.47 [1.33, 1.62] |
| Standing height [z-score] | 0.91 [0.19, 1.59] | 0.51 [-0.26, 1.26] | 0.72 [-0.14, 1.30] | 0.41 [-0.11, 1.13] | 0.63 [-0.07, 1.33] |
| Body mass [kg] | 24.8 [23.0, 28.2] | 31.9 [27.5, 38.0] | 44.9 [39.4, 51.4] | 57.3 [53.1, 62.6] | 38.6 [28.1, 50.4] |
| Leg length [m] | 0.61 [0.57, 0.68] | 0.68 [0.65, 0.73] | 0.77 [0.73, 0.81] | 0.82 [0.79, 0.85] | 0.77 [0.71, 0.82] |
| BMI [kg/m<sup>2</sup>] | 15.9 [14.9, 17.3] | 16.5 [15.1, 18.6] | 18.2 [16.6, 19.9] | 20.2 [18.3, 22.2] | 17.4 [15.8, 19.8] |
| BMI [z-score] | 0.3 [-0.4, 1.1] | 0.1 [-0.8, 1.0] | -0.1 [-0.9, 0.8] | 0.1 [-0.5, 0.8] | 0.09 [-0.7, 0.9] |

BMI = body mass index

### Reference values for the STS

To define the reference values for the STS in children and adolescents, we calculated the distribution of the STS performance for each sex and age group (Table 2 and Figure 1). The median number of repetitions in 1 minute in males (females) ranged from 55 (53) in 14-16 year olds to 61 (64) in 8-10 year olds. In most age groups, 50% of children achieved between 55 and 65 repetitions. No relevant differences in distribution of the number of repetitions were observed between the two genders. The age group 14-16 years showed the lowest median number of repetitions (females 53, males 55).

**Table 2.**
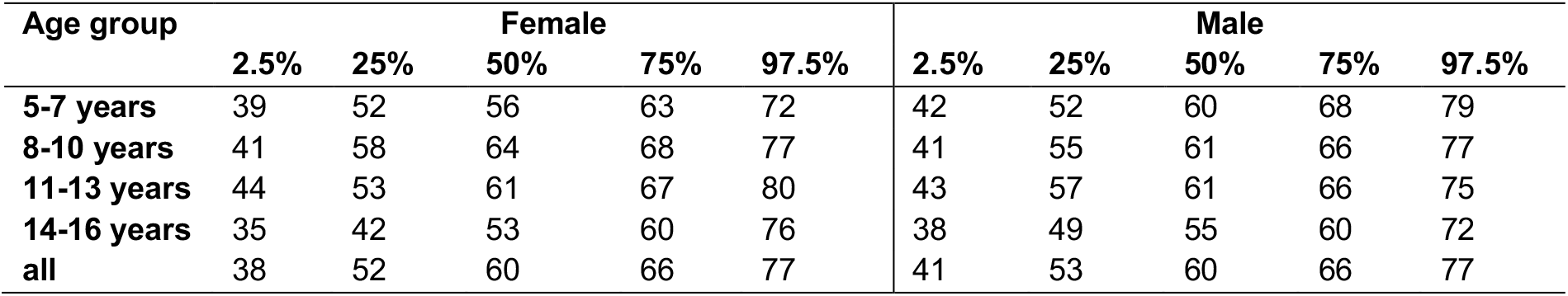
Reference values of the sit-to-stand test in children and adolescents (N = 547)

| Age group | Female |  |  |  |  | Male |  |  |  |  |
| --- | --- | --- | --- | --- | --- | --- | --- | --- | --- | --- |
|  | 2.5% | 25% | 50% | 75% | 97.5% | 2.5% | 25% | 50% | 75% | 97.5% |
| 5-7 years | 39 | 52 | 56 | 63 | 72 | 42 | 52 | 60 | 68 | 79 |
| 8-10 years | 41 | 58 | 64 | 68 | 77 | 41 | 55 | 61 | 66 | 77 |
| 11-13 years | 44 | 53 | 61 | 67 | 80 | 43 | 57 | 61 | 66 | 75 |
| 14-16 years | 35 | 42 | 53 | 60 | 76 | 38 | 49 | 55 | 60 | 72 |
| all | 38 | 52 | 60 | 66 | 77 | 41 | 53 | 60 | 66 | 77 |

**Fig. 1.**
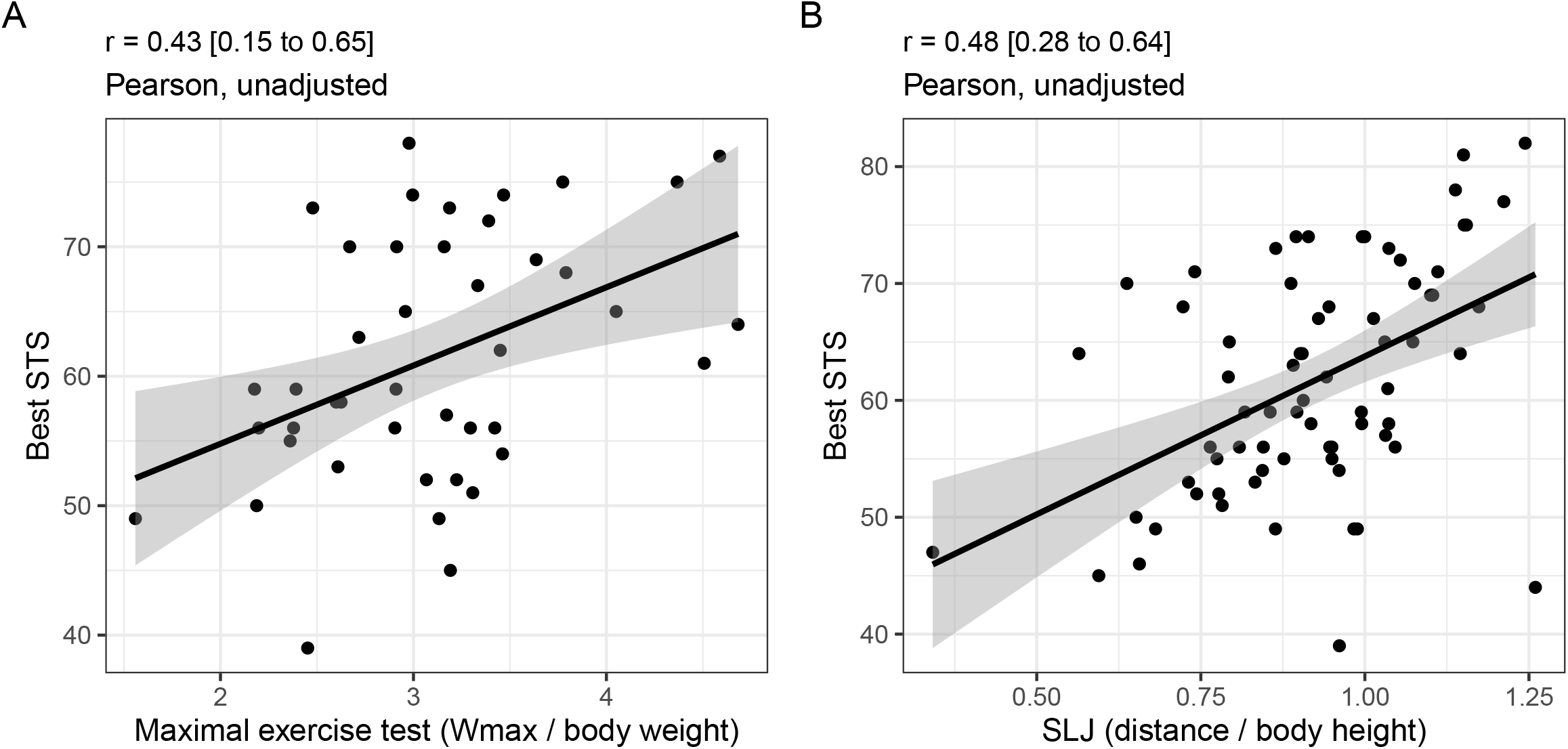
Comparison of STS reference values (median, lines to Q1 and Q3, points for 2.5% and 97.5%) in children and adults (* as published in Strassmann et al. [2]). Reference values in children are calculated based on the best of 2 STS tests, while for adults they are calculated based on a single STS.

We included the best STS performance for the calculation of these reference values. Considering different definitions of STS (first measurement, last measurement, best of two measurements or mean of two repetitions) altered the reference values by approximately 3-5 repetitions (Supplementary Table S2), with those including only the first measurement being lower than the other definitions.

### Reliability

Figure 2 shows the correlation (A) between the results of the two STS tests whereas the Bland-Altman plot (B) shows the mean difference between the two test results as well as the distribution of the differences compared to the number of average repetitions. The second STS had on average 4.8 more repetitions than the first STS (95% limits of agreement −6.7 to 16.4), indicating a clear learning effect.

**Fig. 2.**
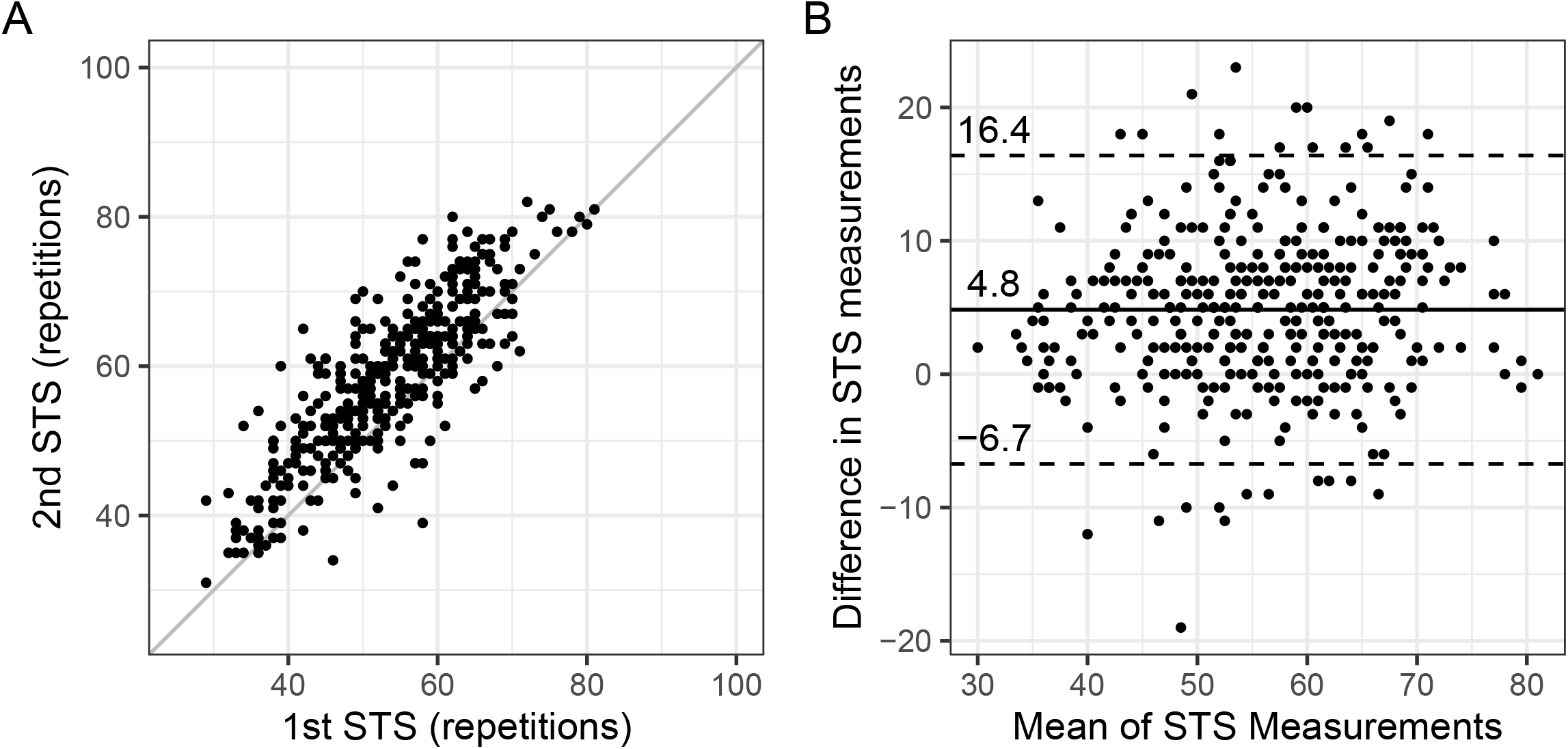
(A) Scatterplot showing the paired results of the first and the second sit-to-stand test (n = 373, diagonal line indicates perfect agreement), (B) Bland-Altman plot showing the mean bias and the limits of agreement (LOA) of the difference between the second and the first sit-to-stand test.

### Construct validity

The relationship between the number of repetitions in the STS and the maximal exercise test and SLJ performance is illustrated in Figure 3. The correlation between STS and aerobic capacity (watt per kilogram body mass) was *r* = 0.43 [95% CI 0.15 to 0.65] and SLJ (distance divided by standing height) *r* = 0.48 [95% CI 0.28 to 0.64].

**Fig. 3.**
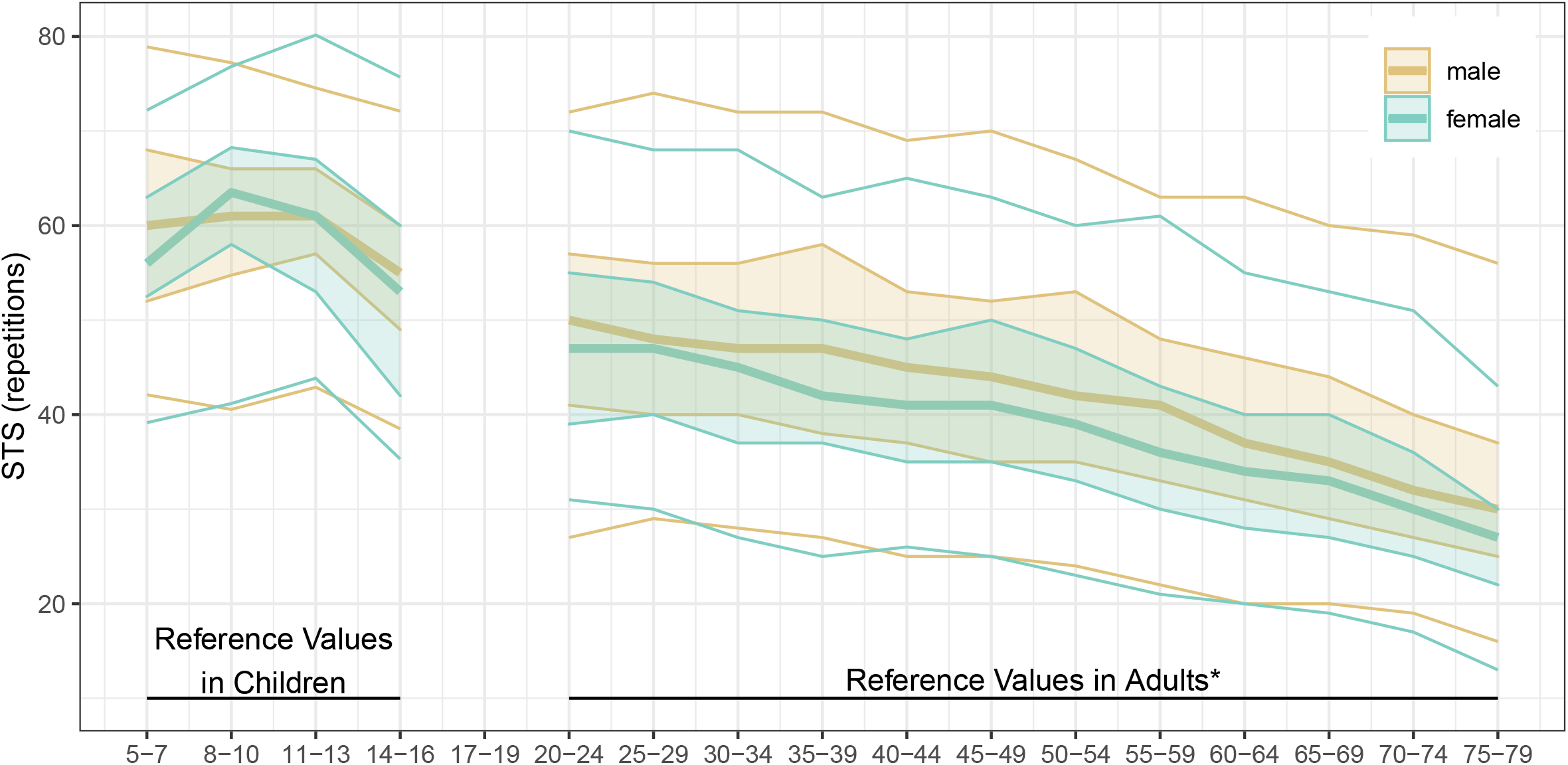
Scatterplot showing the correlation between the mean of the best sit-to-stand (STS) test result and (A) the maximal incremental exercise test on a stationary bike (n = 43), and (B) the standing long jump test (SLJ) (n = 72).

While SLJ and maximal aerobic capacity both increased with age (*r* = 0.44 SLJ, *r* = 0.39 Wmax), STS decreased slightly with age (*r* = −0.14) (Supplementary Figure S1). When adjusting for age using partial correlation, the association was weak to moderate: maximal aerobic exercise test and STS, correlation *r* = 0.36 [95% CI 0.07 to 0.60]; SLJ and STS *r* = 0.42 [95% CI 0.20 to 0.59] (Supplementary Figure S2).

## Discussion

This study provides age and sex-stratified reference values for the 1-minute sit-to-stand test based on the results of a large sample (n = 547) of 5-16 year old healthy children and adolescents which can be used to interpret STS performance in this population. Reference values are necessary to correctly interpret an individual’s STS performance, and to identify individuals with decreased exercise capacity. While there were no significant differences between male and female participants, 14-16 year olds generally performed less well than other age groups. Short-term test-retest reliability was high although there was a clear learning effect of approximately 5 repetitions out of a total range of 40-80 repetitions. STS showed a weak to moderate association with maximal aerobic exercise capacity and lower limb muscle strength.

Concerning construct validity, our study showed only a weak correlation between STS and maximal exercise capacity, in contrast with other studies examining subjects with cystic fibrosis [7, 8, 23]. A low correlation was also observed between STS and SLJ, as a proxy for lower limb muscle power, and between STS and body mass index (BMI z-scores), as a proxy for muscle mass.

The main strengths of this study are the large sample size (n = 547), to our knowledge the largest study to date of STS in healthy children and adolescents, as well as the consideration of different age groups. A limitation of this study is that it is based on a convenience sample, not a population-based sample, and that the sample on which the examination of construct validity is based is not a random subsample. A learning effect could only be evaluated in a subpopulation of children (n = 373, 68%) for which two STS tests were available. However, computing reference values for STS based on this subset had only a small impact on the STS percentiles (i.e., differences of min −2 to max 2 repetitions across age groups and sexes). Additionally, the maximal exercise test and SLJ used to assess construct validity were not measured in all subjects.

There have been few other studies of STS in children. A recent study in Belgium [12] assessed construct validity and test-retest reliability of STS in a sample of 52 children aged 8-18 years, comparing it to the 6MWT and final heart rate. Similarly to our study, they found a learning effect in the number of STS repetitions between the first and second trial (mean difference 2.5 repetitions, 95% CI 1.5 to 3.5), suggesting that at least one practice test is needed to ensure an accurate estimate of functional exercise capacity. The same research group also developed a predictive model for STS in healthy 6 to 12 year olds (n = 238) [13], focusing on individual characteristics (age, sex, height and weight) and final heart rate. They reported differing mean repetitions of 47 in girls and 54 in boys, while we observed the same median of 61 in both boys and girls in this age range. Gender differences may have occurred due to only two different chair heights (46cm chair for participants 140cm or taller, otherwise 32cm chair) instead of individually adapted height as in our study (median 40cm, range 24 to 50.5cm), that may have induced an advantage of one gender or a disadvantage for the other in a rather small and heterogeneous sample with a large age range. Our study is the largest study of STS in children to date and is the only to provide reference values for a healthy population of children and adolescents in different age categories.

We provided reference values for STS in healthy participants aged 6 to 16. Although there were no differences between males and females, older participants (14-16 years) performed less well on average than those in younger age groups, even though muscle mass and strength are still increasing at this age. We suspected that weaker performance of this age group might be related to greater height or longer leg length, but only weak correlations were observed (Supplementary Figure S3) between STS and height (*r* = −0.16, 95% CI −0.24 to −0.08) or leg length (*r* = −0.13, 95% CI −0.24 to − 0.02) or ratio of leg length to height (*r* = 0.09, 95% CI −0.02 to 0.21). Adjusting for age using partial correlation did not increase the strength of the associations between STS and height (*r* = −0.10, 95% CI −0.18 to −0.02, adjusted for age) or leg length (*r* = 0.02, 95% CI −0.10 to 0.13, adjusted for age) or the ratio of leg length to height (*r* = 0.08, 95% CI −0.04 to 0.19, adjusted for age). It cannot be excluded that the age differences in STS performance were due to lack of motivation or decreased physical activity commonly found during adolescence [24]. However, it could also be that performance in STS reaches its peak by the age of 11 and then decreases in accordance to physical activity levels [25]. This hypothesis would be consistent with the findings of a study of STS in adults, which reported STS values for 20-24 year olds which were lower than what we observed in 14-16 year olds, indicating that STS performance decreases steadily with age (Figure 1) [2]. However, as there is no data available for 17-19 year olds, it would be necessary to include such participants in future studies to verify this hypothesis.

Regarding construct validity, the weak to moderate correlation we observed between STS and maximal exercise capacity (Wmax) suggests that neither maximal nor submaximal [12] aerobic capacity may not be one of the major determinants of exercise in health children and adolescents, and therefore it is possible that STS is not best suited to assess aerobic capacity in this population. Correlation with SLJ as a measure of lower limb muscle power was also weak, despite STS having been shown previously to be moderately associated with muscle strength in COPD patients [1]. In a predominantly normal weight population as described here, one would suggest that BMI serves as a proxy measure for muscle mass, and would therefore be related to STS. However, only a weak correlation was observed here between BMI and STS. The prediction model proposed by Reychler et al. [13] suggests that sex, age, weight, and final heart rate explain approximately 24% of STS performance. Taken together, these findings suggest that the STS can be taken neither as an aerobic performance test, nor as a pure strength test, as has been observed in other studies [1], but may also include skills like balance and coordination.

The results of this study suggest various avenues for future research. First, a systematic review of STS should be undertaken to determine the current state of evidence in both children and adults. Second, technical standards for proper conduct of STS are needed, especially for use in children (e.g., where a standard size chair may be too large). Third, further studies are needed to evaluate the learning effect over multiple STS measurements, to provide standard values for other racial or ethnic groups and in other geographic regions, and to better validate STS against other measures of physical fitness such as anaerobic exercise capacity measured by a Wingate anaerobic test [26, 27] or steep ramp test [28]. Such a study should also include 17 to 19 year olds as data on subjects in this age group are missing in both the present study and in the previously published [2] reference values for STS in adults.

## Conclusions

This study provides reference values for the STS in healthy children and adolescents aged 5-16 years. Reference values help to interpret the performances achieved in STS and to detect individuals with decreased physical performance. Further investigation is needed to clarify whether the STS primarily demands muscle strength or aerobic capacity in children and adolescents.

## Data Availability

Data may be available upon reasons request to the corresponding author.

## Compliance with Ethical Standards

### Funding source

No sources of funding were used to assist in the preparation on this article.

### Patient and Public Involvement statement

Participants were not involved in the design, recruitment, or conduct of this study.

### Availability of data and material

Data may be available upon reasons request to the corresponding author.

### Code availability

The R script is available in Supplementary Material 2.

## Author’s contributions

SH, SK, and TR: made substantial contributions to conception and design; JS, TF, SK, and TR: contributed to data collection; SH carried out data analysis; SH, JS, UG, SK and TR: interpreted the data; SH and TF: wrote the first draft of the manuscript and all authors were involved in revising it critically for important intellectual content; all authors provide final approval of the version to be published and agreed to be accountable for all aspects of the work.

