## Supplemental File 1 for "Reference values and validation of the 1-min sit-to-stand test in healthy 5- to 16-year-old youth: a cross-sectional study"

*Electronic Supplementary Material Appendix*

Sarah R Haile, Thea Führer, Urs Granacher, Julien Stocker, Thomas Radtke, Susi Kriemler

**Table S1** Participation in the various physical fitness test by sex and age group

| sex | age group | n | STS | SLJ | CPET |
|---|---|---|---|---|---|
| Male | 5-7 years | 63 | 73% | 38% | 16% |
| Male | 8-10 years | 72 | 46% | 10% | 7% |
| Male | 11-13 years | 90 | 72% | 4% | 4% |
| Male | 14-16 years | 53 | 92% | 8% | 8% |
| Female | 5-7 years | 59 | 64% | 34% | 20% |
| Female | 8-10 years | 88 | 50% | 6% | 5% |
| Female | 11-13 years | 75 | 75% | 8% | 5% |
| Female | 14-16 years | 47 | 89% | 4% | 0% |
| all | all | 547 | 68% | 13% | 8% |

**Table S2** Reference values (median [Q1 to Q3], by sex, age group and definition of STS (first measurement, last measurement, best measurement or mean repetitions)

| sex | age group | first | last | best | mean |
|---|---|---|---|---|---|
| Male | 5-7 years | 56 [48 to 62] | 60 [52 to 68] | 60 [52 to 68] | 58 [51 to 65] |
| Male | 8-10 years | 59 [50 to 64] | 61 [55 to 66] | 61 [55 to 66] | 60 [53 to 66] |
| Male | 11-13 years | 58 [51 to 63] | 61 [56 to 66] | 61 [57 to 66] | 59 [54 to 64] |
| Male | 14-16 years | 49 [44 to 55] | 54 [49 to 60] | 55 [49 to 60] | 52 [46 to 56] |
| Female | 5-7 years | 53 [48 to 59] | 56 [52 to 62] | 56 [52 to 63] | 54 [51 to 60] |
| Female | 8-10 years | 60 [55 to 66] | 63 [57 to 67] | 64 [58 to 68] | 62 [57 to 67] |
| Female | 11-13 years | 58 [50 to 64] | 61 [53 to 66] | 61 [53 to 67] | 60 [51 to 64] |
| Female | 14-16 years | 47 [38 to 56] | 53 [42 to 60] | 53 [42 to 60] | 50 [40 to 58] |

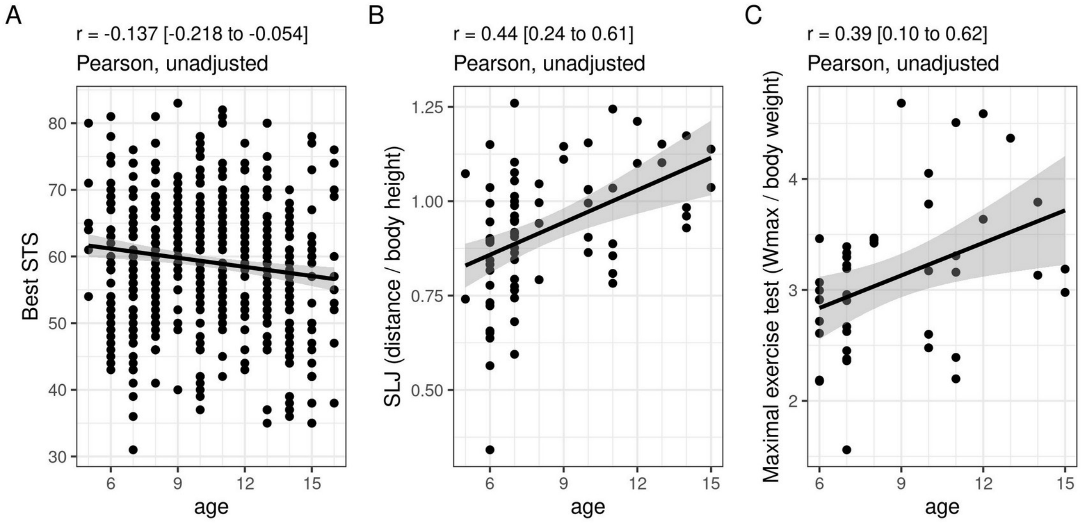


**Figure S1** Association of three key measures (A) = sit-to-stand test (STS), (B) = standing long jump test (SLJ / body height), and (C) = cardiopulmonary exercise maximal incremental exercise test on a stationary bike (Wmax / body weight) with age

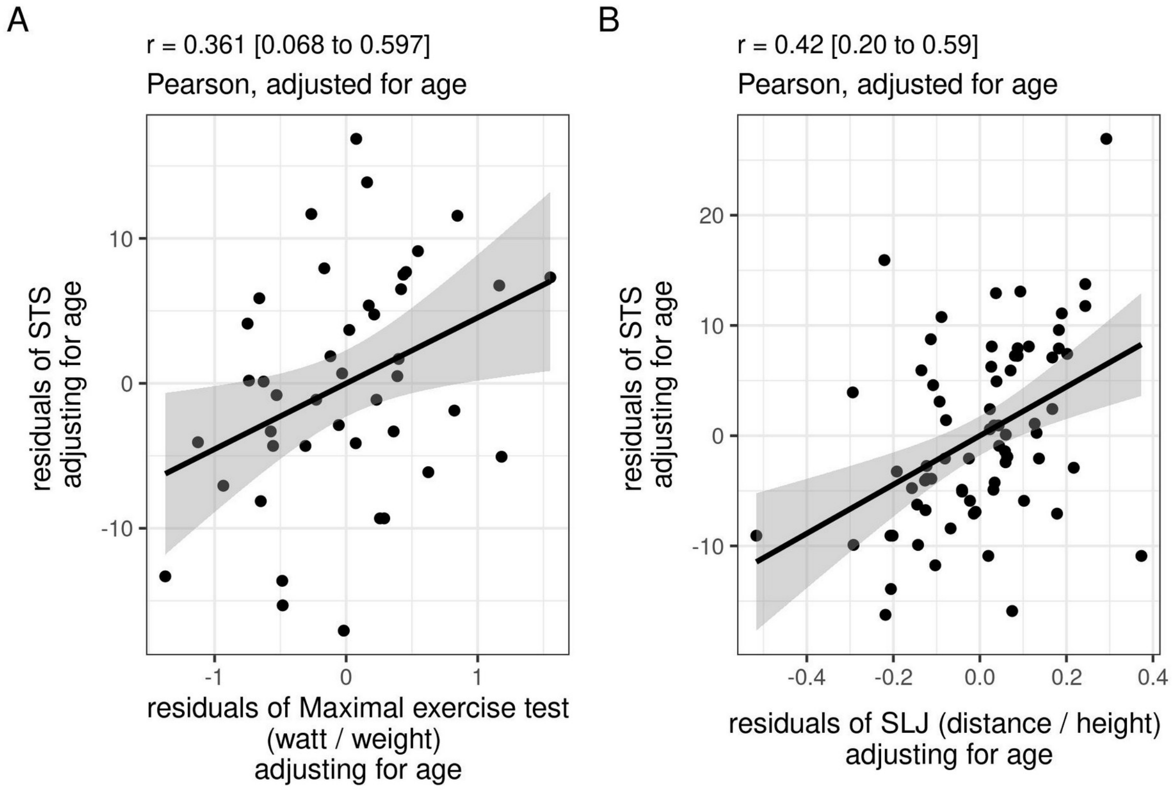


**Figure S2** Scatterplot showing the correlation between the mean of the two sit-to-stand test (STS) results and (A) the maximal incremental exercise test on a stationary bike (Wmax), and (B) the standing long jump test (SLJ), adjusting for age

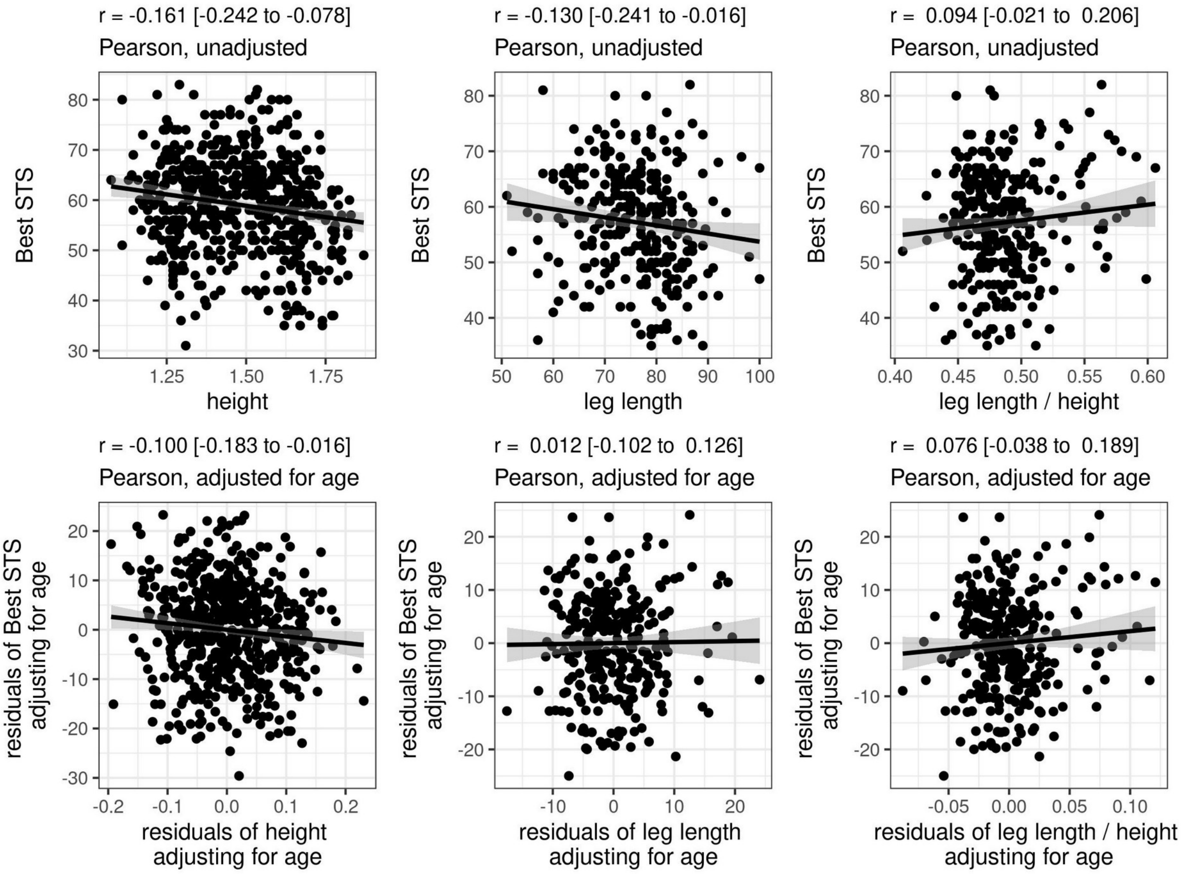


**Figure S3** Association of STS with height (left), leg length (middle), and ratio of leg length to height (right), both unadjusted (top row), and adjusted for age (bottom row)

**Working instructions - 1-minute Sit-to-Stand**

**Equipment**

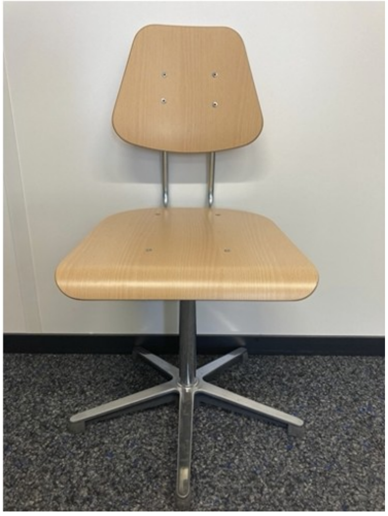

- Height adjustable chair
- Tape measure
- Stopwatch/Smartphone
- Counter (compulsory)
- Heart rate monitor (compulsory)
- Pulse oximeter (compulsory, for chronic disease populations)
- Scales to assess leg fatigue and breathlessness (e.g., Borg Scale)
- Solid footwear/sturdy shoes

**Pre-test preparations**
- Place the chair against the wall for safety reasons.
- Adjust the seat height to have approximately 90° knee angle flexion.
- Due to the observed learning effect, it is important that the child performs <u>at least one practice test</u>. If two tests are performed on the same day, consider a rest of about 10 minutes [1] between the first and second test to allow for sufficient recovery.

**Test instructions**
- Children are asked to perform as <u>many repetitions as possible</u> in 1 minute.
- Instruct the child to sit with his/her legs hip-width apart and with the hands placed on the hips.
- Children are not allowed to use their hands or arms to assist movement.
- Start position: Children sitting on a chair.
- Instruct the child to stand completely straight and to touch the chair with his/her bottom when sitting. They don’t need to sit fully back on the chair. Correct during the test, if needed. Only correctly performed STS repetitions are counted.
- During the test, children are allowed to have a rest, when needed.
- Do <u>not</u> verbally encourage the child during the test. After 45 s the child is told that “15 s left until the test is over”.

**Outcomes**
- Number of repetitions
- Heart rate
- Oxygen saturation (chronic disease populations)
- Subjective perception of breathlessness and leg fatigue
- STS Power Index (compulsory)

**Reference values** (N = 547)

| | Female | | | | | Male | | | | |
|---|---|---|---|---|---|---|---|---|---|---|
| **Age** | **2.5%** | **25%** | **50%** | **75%** | **97.5%** | **2.5%** | **25%** | **50%** | **75%** | **97.5%** |
| **5-7 y** | 39 | 52 | 56 | 63 | 72 | 42 | 52 | 60 | 68 | 79 |
| **8-10 y** | 41 | 58 | 64 | 68 | 77 | 41 | 55 | 61 | 66 | 77 |
| **11-13 y** | 44 | 53 | 61 | 67 | 80 | 43 | 57 | 61 | 66 | 75 |
| **14-16 y** | 35 | 42 | 53 | 60 | 76 | 38 | 49 | 55 | 60 | 72 |
| **all** | 38 | 52 | 60 | 66 | 77 | 41 | 53 | 60 | 66 | 77 |
