## Supplemental File 2 for "Reference values and validation of the 1-min sit-to-stand test in healthy 5- to 16-year-old youth: a cross-sectional study"

### Reference values and Validation of the 1-min sit-to-stand test in children and adolescents - results

2021-03-23

```
## # A tibble: 5 x 5
## # Groups:   excl_age, excl_miss_age, excl_miss_sts [5]
##   excl_age excl_miss_age excl_miss_sts excl_motivation     n
##      <dbl>         <dbl>         <dbl>           <dbl> <int>
## 1        0             0             0               1    19
## 2        0             0             1               0     2
## 3        0             1             0               0     5
## 4        0             1             1               0     1
## 5        1             0             0               0    13
```

##### 0.1 Statistical Analysis

Key baseline variables have been summarized as n (%) or median [interquartile range], by sex and age group (5-7, 8-10, 11-13, 14-16). Reference values for STS are reported using the following percentiles, by age group and sex: 2.5, 25, 50, 75, 97.5. Reliability between first and second STS tests was assessed using the method of [1], where mean difference in the two tests assesses agreement between the two measurements, along with 95% limits of agreement. Construct validity was examined by comparing STS to standing long jump and the bicycle test using Pearson correlation. We also consider correlation of the three measures with body height, body weight and leg length, adjusting for age. Correlation coefficients of 0-0.19 were considered very weak, 0.2-0.39 weak, 0.4-0.59 moderate, 0.60-0.79 strong and 0.80–1.0 as very strong [2]. Analysis was performed using the R programming language [3] (R Version 4.0.3).

#### 1 Results

##### 1.1 Study Population

Table 1: Summary statistics for all data (including all participants 5-16 with at least 1 STS)

| name | 5-7 | 8-10 | 11-13 | 14-16 | all |
|---|---|---|---|---|---|
| n | 122 | 160 | 165 | 100 | 547 |
| sex_f | 59 (48.4%) | 88 (55%) | 75 (45.5%) | 47 (47%) | 269 (49.2%) |
| ht | 1.26 [1.22, 1.30] | 1.39 [1.34, 1.45] | 1.57 [1.52, 1.64] | 1.67 [1.63, 1.74] | 1.47 [1.33, 1.62] |
| ht_z | 0.91 [0.19, 1.59] | 0.51 [-0.26, 1.26] | 0.72 [-0.14, 1.30] | 0.41 [-0.11, 1.13] | 0.63 [-0.072, 1.33] |
| wt | 24.8 [23.0, 28.2] | 31.9 [27.5, 38.0] | 44.9 [39.4, 51.4] | 57.3 [53.1, 62.6] | 38.6 [28.1, 50.4] |
| leg | 0.610 [0.570, 0.675] | 0.680 [0.645, 0.730] | 0.770 [0.730, 0.810] | 0.820 [0.790, 0.850] | 0.770 [0.710, 0.820] |
| chair | 33.0 [31.0, 34.0] | 36.1 [35.0, 38.1] | 42.0 [40.0, 43.4] | 44.0 [43.0, 46.0] | 40.0 [35.0, 43.0] |
| BMI | 15.9 [14.9, 17.3] | 16.5 [15.1, 18.5] | 18.2 [16.6, 19.9] | 20.2 [18.3, 22.2] | 17.4 [15.8, 19.8] |
| BMI_z | 0.3 [-0.4, 1.1] | 0.1 [-0.8, 1.0] | -0.1 [-0.9, 0.8] | 0.1 [-0.5, 0.8] | 0.09 [-0.7, 0.9] |

```
##    Min. 1st Qu.  Median    Mean 3rd Qu.    Max.    NA's 
##   24.00   35.00   40.00   39.25   43.00   50.50      59 
```

#### 1.2 Reference values for the STS test

Table 2: Reference values of the sit-to-stand test in children and adolescents (N = 547)

| agegrp | F_p025 | F_p25 | F_p50 | F_p75 | F_p975 | M_p025 | M_p25 | M_p50 | M_p75 | M_p975 |
|---|---|---|---|---|---|---|---|---|---|---|
| 5-7 | 39 | 52 | 56 | 63 | 72 | 42 | 52 | 60 | 68 | 79 |
| 8-10 | 41 | 58 | 64 | 68 | 77 | 41 | 55 | 61 | 66 | 77 |
| 11-13 | 44 | 53 | 61 | 67 | 80 | 43 | 57 | 61 | 66 | 75 |
| 14-16 | 35 | 42 | 53 | 60 | 76 | 38 | 49 | 55 | 60 | 72 |
| all | 38 | 52 | 60 | 66 | 77 | 41 | 53 | 60 | 66 | 77 |

```
## # A tibble: 2 x 8
##   sex       n sts_mean sts_p025 sts_q1 sts_med sts_q3 sts_p975
##   <fct> <int>    <dbl>    <dbl>  <dbl>   <dbl>  <dbl>    <dbl>
## 1 m       184     60.4     41.6     55      61     66     77  
## 2 f       194     60.5     41.8     55      61     67     77.2
```

#### 1.3 Reliability

The second STS had on average 4.8 more repetitions than the first STS (95% limits of agreement -6.7 to 16.4), indicating a possible learning effect.

#### 1.4 Construct validity

```
## [1] 0.4798711
## [1] 0.431893
```

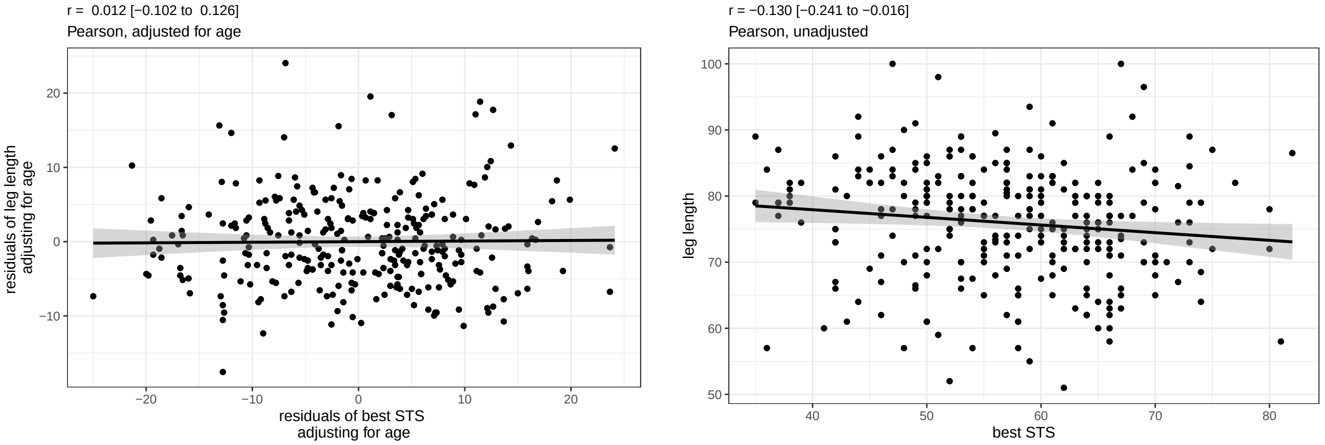

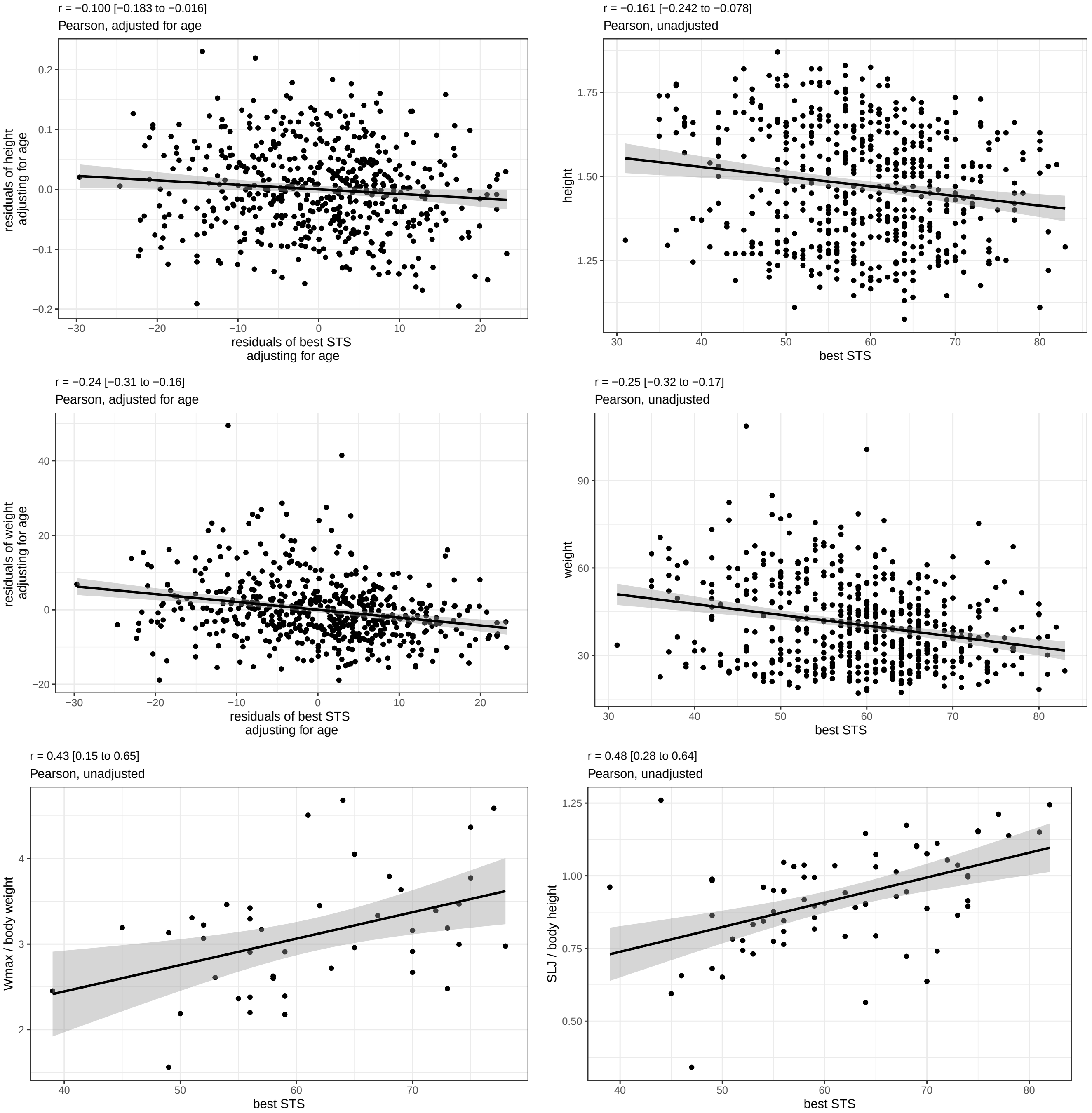

### Figures

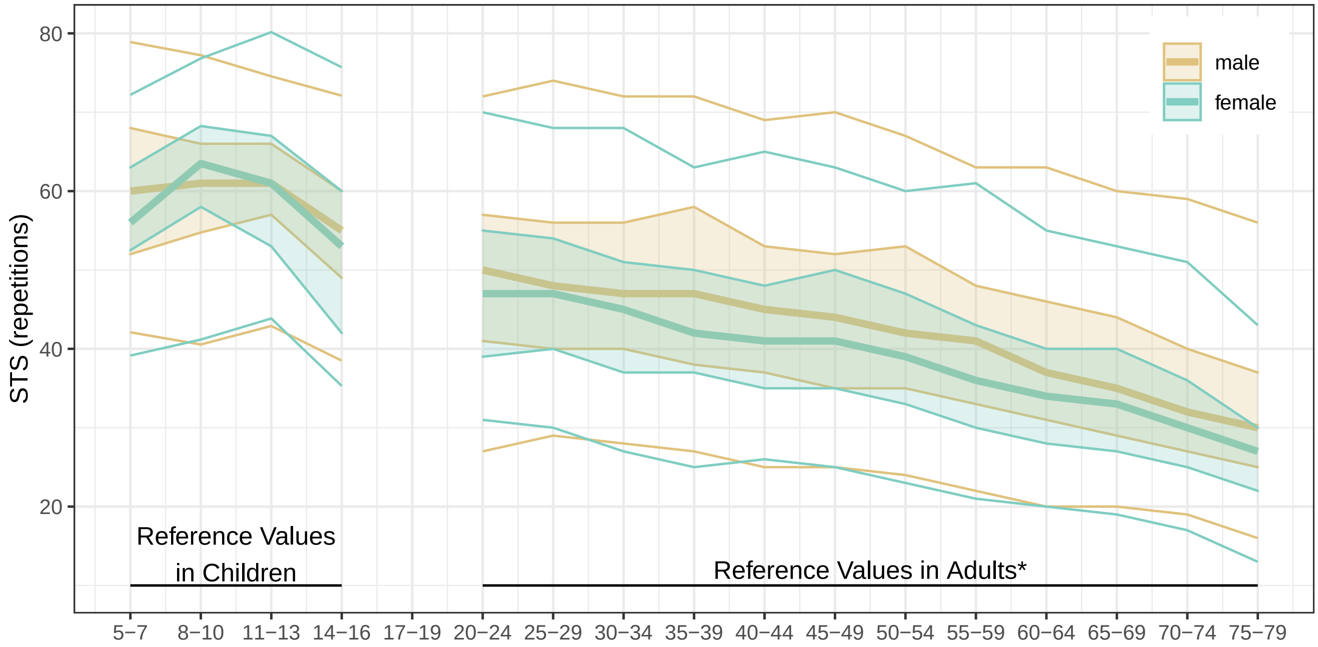


Figure 1: Comparison of STS reference values (median, lines to Q1 and Q3, points for 2.5% and 97.5%) in children and adults (* as published in Strassman et al 2013)

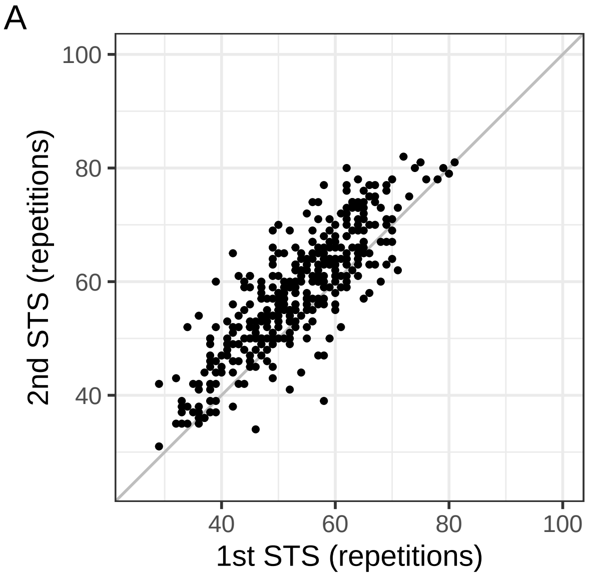


Figure 2: (A) Scatterplot showing the paired results of the first and the second sit-to-stand test (diagonal line indicates perfect agreement), (B) Bland-Altman plot showing the mean bias and the limits of agreement (LOA) of the difference between the second and the first sit-to-stand test

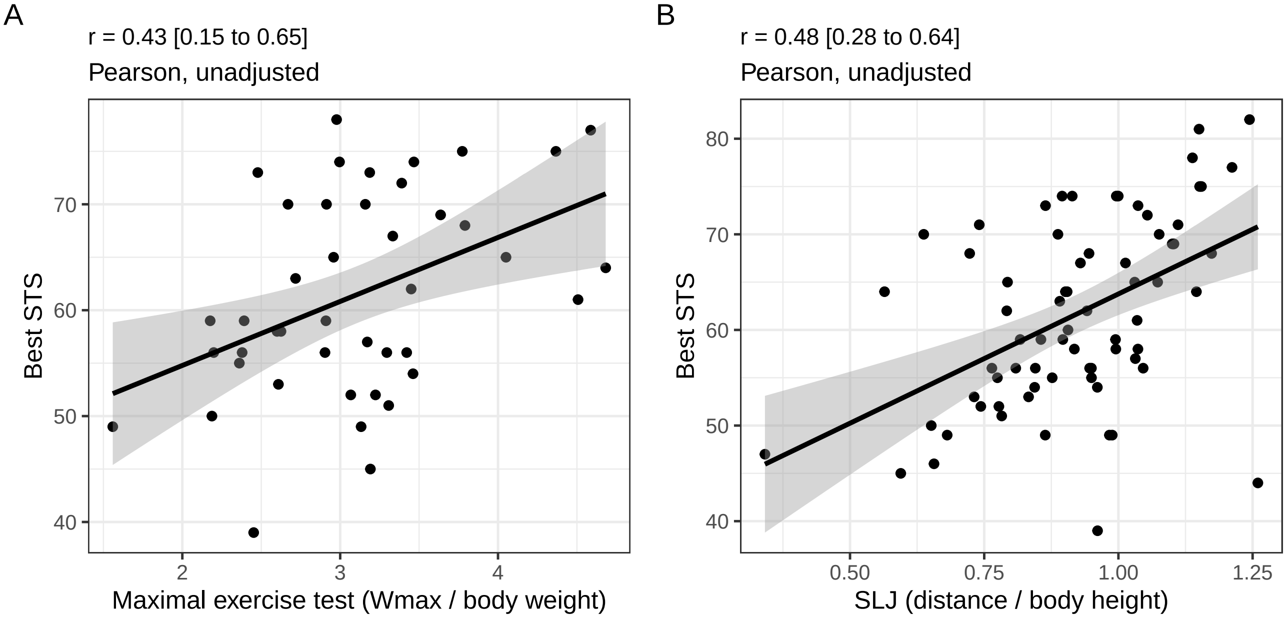


Figure 3: Scatterplot showing the correlation between the mean of the two sit-to-stand test results and A) the bicycle test, and B) the standing long jump.

### Supplementary Material

Table S1: Participation in other measures of physical fitness by age and sex

| sex | agegrp | n | p_STS_2 | p_SLJ | p_Wmax |
|---|---|---|---|---|---|
| m | 5-7 | 63 | 73% | 38% | 16% |
| m | 8-10 | 72 | 46% | 10% | 7% |
| m | 11-13 | 90 | 72% | 4% | 4% |
| m | 14-16 | 53 | 92% | 8% | 8% |
| f | 5-7 | 59 | 64% | 34% | 20% |
| f | 8-10 | 88 | 50% | 6% | 5% |
| f | 11-13 | 75 | 75% | 8% | 5% |
| f | 14-16 | 47 | 89% | 4% | 0% |
| all | all | 547 | 68% | 13% | 8% |

Table S2: Reference values (median [Q1 to Q3], by sex, age group and definition of STS

| sex | agegrp | first | best | last | mean |
|---|---|---|---|---|---|
| m | 5-7 | 56 [48 to 62] | 60 [52 to 68] | 60 [52 to 68] | 58 [51 to 65] |
| m | 8-10 | 59 [50 to 64] | 61 [55 to 66] | 61 [55 to 66] | 60 [53 to 66] |
| m | 11-13 | 58 [51 to 63] | 61 [57 to 66] | 61 [56 to 66] | 59 [54 to 64] |
| m | 14-16 | 49 [44 to 55] | 55 [49 to 60] | 54 [49 to 60] | 52 [46 to 56] |
| f | 5-7 | 53 [48 to 59] | 56 [52 to 63] | 56 [52 to 62] | 54 [51 to 60] |
| f | 8-10 | 60 [55 to 66] | 64 [58 to 68] | 63 [57 to 67] | 62 [57 to 67] |
| f | 11-13 | 58 [50 to 64] | 61 [53 to 67] | 61 [53 to 66] | 60 [51 to 64] |
| f | 14-16 | 47 [38 to 56] | 53 [42 to 60] | 53 [42 to 60] | 50 [40 to 58] |

```
## # A tibble: 16 x 7
## # Groups:   sex, agegrp [8]
##    sex   agegrp definition subset   p50    q1    q3
##    <fct> <fct>  <chr>      <chr>  <dbl> <dbl> <dbl>
##  1 m     5-7    best       all    60    52    68
##  2 m     5-7    best       both   61    52    69
##  3 m     8-10   best       all    61    54.8  66
##  4 m     8-10   best       both   62    57    64
##  5 m     11-13  best       all    61    57    66
##  6 m     11-13  best       both   61    55    66
##  7 m     14-16  best       all    55    49    60
##  8 m     14-16  best       both   55    49    60
##  9 f     5-7    best       all    56    52.5  63
## 10 f     5-7    best       both   56    53.2  63
## 11 f     8-10   best       all    63.5  58    68.2
## 12 f     8-10   best       both   65.5  59.5  71
## 13 f     11-13  best       all    61    53    67
## 14 f     11-13  best       both   59.5  52    66.2
## 15 f     14-16  best       all    53    42    60
## 16 f     14-16  best       both   51    42    59
```

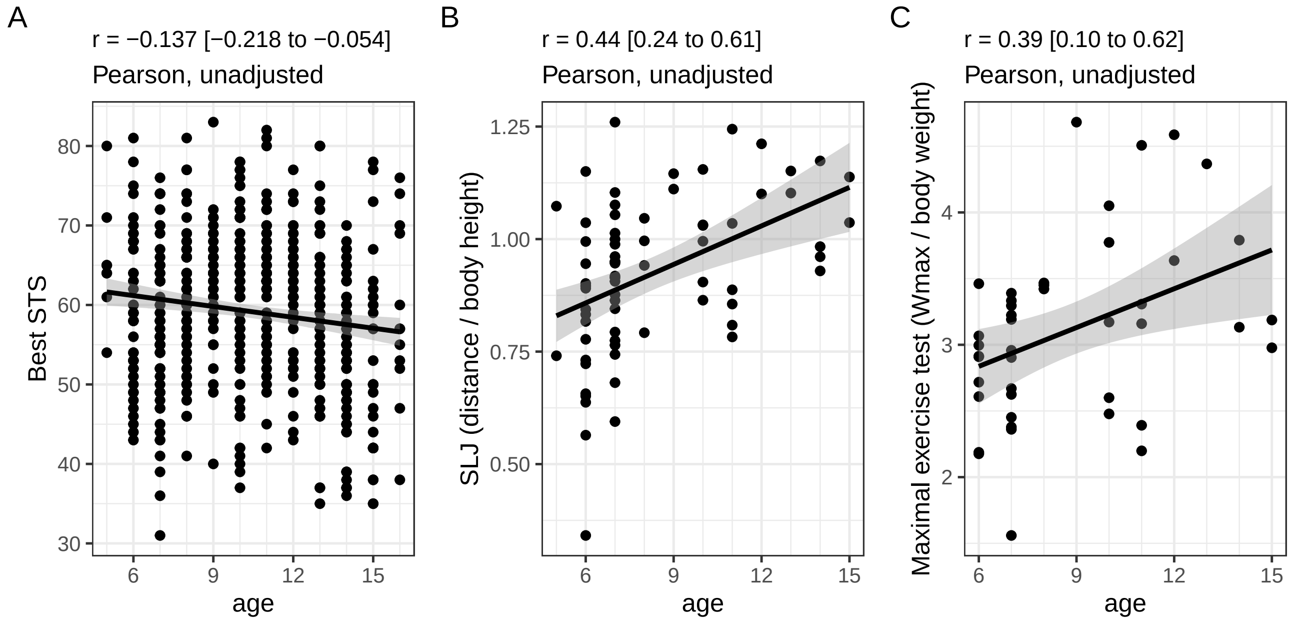


Figure S1: Association of three key measures (STS, SLJ and maximal exercise test) with age

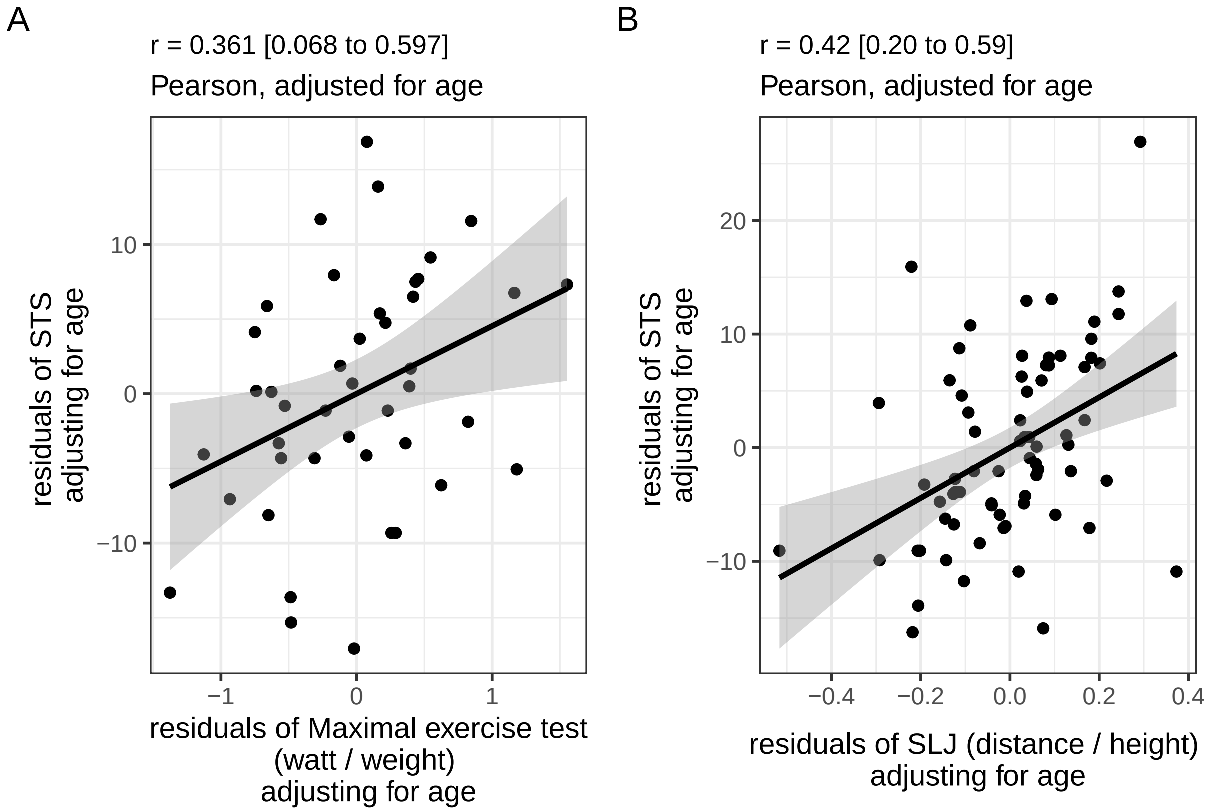


Figure S2: Scatterplot showing the correlation between the 1st the two sit-to-stand test results and A) the bicycle test, and B) the standing long jump, adjusting for age

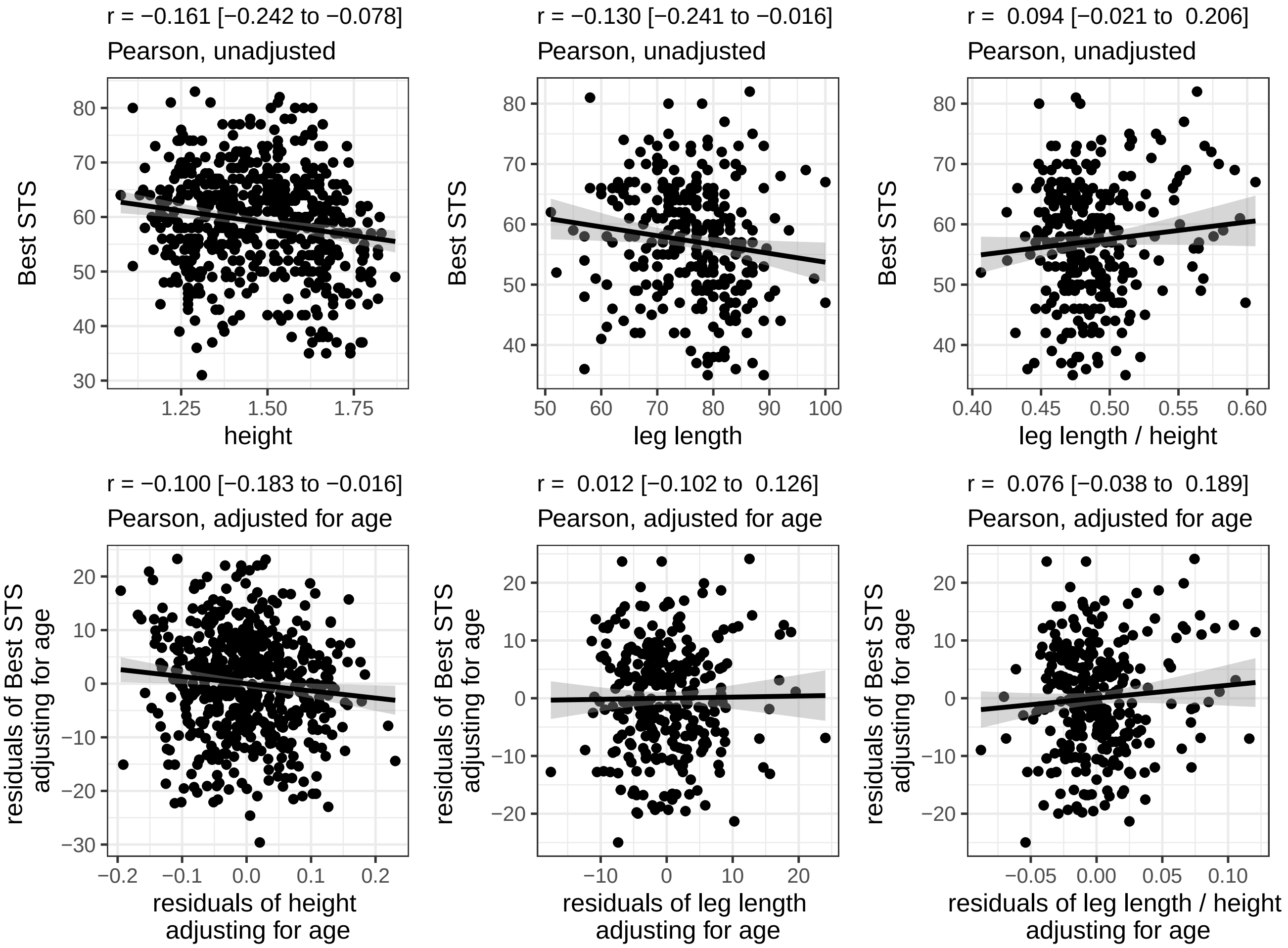


Figure S3: Association of STS with height, leg length, and ratio of leg length to height

#### Code Appendix

```
knitr::opts_chunk$set(echo = FALSE, warning = FALSE, message = FALSE, dev = c('pdf', "jpeg", "postscript")
options(knit.r.table.format = "markdown")
options(scipen = 8, show.signif.stars = FALSE)
library(tidyverse)
theme_set(theme_bw())
library(readxl)
library(broom)
library(tableone)
library(patchwork)
datfile <- "STS Datensatz alle 20200316.xlsx"

zurich <- read_xlsx(datfile, sheet = 1)
zurich  <- zurich %>%
  select(id = ID_Kind,
         source = Datenherkunft,
         age = "Alter (in Jahren)",
         sex = "Geschlecht 1=m 2=f",
         wt = "Gewicht (kg)",
         ht =  "Grösse (m)",
         leg = "Beinlänge (cm)",
         chair = "Stuhlhöhe (cm)",
         sts1 = "STS Rep 1",
         sts2 = "STS Rep 2",
         shuttle = "Shuttle Run (min)",
         Bemerkungen,
         Zusatzinformation,
         Exclude) %>%
  mutate_at(vars(age:shuttle), as.numeric) %>%
  mutate(id = as.character(id)) %>%
  mutate(ht = ifelse(ht > 0, ht, NA))

basel <- read_xlsx(datfile, sheet = 2)
basel <- basel %>%
  select(id = ID_Kind,
         source = Datenherkunft,
         age = "Alter (in Jahren)",
         sex = "Geschlecht 1=m 2=f",
         wt = "Gewicht (kg)",
         ht =  "Körperhöhe (cm)",
         leg = "Beinlänge (cm)",
         chair = "Stuhlhöhe (cm)",
         sts1 = "STS Rep 1",
         sts2 = "STS Rep 2",
         slj1 = "SLJ Rep 1 (m)",
         slj2 = "SLJ  Rep 2 (m)",
         slj3 = "SLJ Rep 3 (m)",
         shuttle = "Shuttle Run (min)",
         watt = "extrapolierte Wattzahl (WATT)") %>%
  mutate_at(vars(wt:watt), as.numeric) %>%
  mutate_at(c("slj1", "slj2", "slj3"), ~ ifelse(. > 5, . / 100, .))
```

```
validdat <- read_xlsx(datfile, sheet = 3)
validdat <- validdat %>%
  select(id = ID_Kind,
         source = Datenherkunft,
         age = "Alter (in Jahren)",
         sex = "Geschlecht 1=m 2=f",
         wt = "Gewicht (kg)",
         ht =  "Körperhöhe (cm)",
         leg = "Beinlänge (cm)",
         chair = "Stuhlhöhe (cm)",
         sts1 = "STS Rep 1",
         sts2 = "STS Rep 2",
         slj1 = "SLJ Rep 1 (m)",
         slj2 = "SLJ  Rep 2 (m)",
         slj3 = "SLJ Rep 3 (m)",
         shuttle = "Shuttle Run (min)",
         watt = "extrapolierte Wattzahl (WATT)",
         maxtest = "Max. Test? 0=ja; 1=nein") %>%
  filter(source %in% c("BSM / UKBB", "SURfit Kids", "Urs / DE")) %>%
  mutate_at(vars(sex:watt), as.numeric) %>%
  mutate(slj2 = ifelse(id == "Test 19", 1.23, slj2),
         maxtest = factor(maxtest, 0:1, c("yes", "no")))

combine2 <- function(x, y){
  if(is.na(x) & is.na(y)){
    out <- NA
  } else if(is.na(x)){
    out <- y
  } else if(is.na(y)){
    out <- x
  } else if(x == y){
    out <- y
  } else if(x != y){
    warning("values don't match")
    out <- paste("ERROR", x, y)
  }
  out
}

zb <- full_join(zurich, basel, by = c("id", "source")) %>%
  mutate(age = map2_dbl(age.x, age.y, combine2),
         sex = map2_dbl(sex.x, sex.y, combine2),
         wt = map2_dbl(wt.x, wt.y, combine2),
         ht = map2_dbl(ht.x, ht.y, combine2),
         leg = map2_dbl(leg.x, leg.y, combine2),
         chair = map2_dbl(chair.x, chair.y, combine2),
         sts1 = map2_dbl(sts1.x, sts1.y, combine2),
         sts2 = map2_dbl(sts2.x, sts2.y, combine2),
         shuttle = map2_dbl(shuttle.x, shuttle.y, combine2)) %>%
  select(id, source, Exclude, Bemerkungen, Zusatzinformation,
         age, sex, ht, wt, leg, chair,
         sts1, sts2, slj1, slj2, slj3, shuttle)
```

```r
alldat <- zb %>% full_join(validdat, by = c("id", "source")) %>%
  mutate(age = map2_dbl(age.x, age.y, combine2),
         sex = map2_dbl(sex.x, sex.y, combine2),
         wt = map2_dbl(wt.x, wt.y, combine2),
         ht = map2_dbl(ht.x, ht.y, combine2),
         leg = map2_dbl(leg.x, leg.y, combine2),
         chair = map2_dbl(chair.x, chair.y, combine2),
         sts1 = map2_dbl(sts1.x, sts1.y, combine2),
         sts2 = map2_dbl(sts2.x, sts2.y, combine2),
         slj1 = map2_dbl(slj1.x, slj1.y, combine2),
         slj2 = map2_dbl(slj2.x, slj2.y, combine2),
         slj3 = map2_dbl(slj3.x, slj3.y, combine2),
         shuttle = map2_dbl(shuttle.x, shuttle.y, combine2)) %>%
  mutate(ht = ifelse(ht == 0, NA, ht)) %>%
  select(id, source, Exclude, Bemerkungen, Zusatzinformation,
         age, sex, ht, wt, leg, chair,
         sts1, sts2, slj1, slj2, slj3, shuttle, watt, maxtest)

keep_vars <- c("id")
excl_age <- alldat %>% filter(!(age >= 5 & age <= 16)) %>%
  select(all_of(keep_vars)) %>% mutate(excl_age = 1)
excl_miss_age <- alldat %>% filter(is.na(age)) %>%
  select(all_of(keep_vars))  %>% mutate(excl_miss_age = 1)
excl_miss_sts <- alldat %>% filter(!(!is.na(sts1) | !is.na(sts2))) %>%
  select(all_of(keep_vars))  %>% mutate(excl_miss_sts = 1)
excl_motivation <- alldat %>% filter(Exclude == 1) %>%
  select(all_of(keep_vars)) %>% mutate(excl_motivation = 1)

excl_all <- full_join(excl_age, excl_miss_age, by = "id") %>%
  full_join(excl_miss_sts, by = "id") %>%
  full_join(excl_motivation, by = "id") %>%
  mutate(excl_age = replace_na(excl_age, 0),
         excl_miss_age = replace_na(excl_miss_age, 0),
         excl_miss_sts = replace_na(excl_miss_sts, 0),
         excl_motivation = replace_na(excl_motivation, 0)) %>%
  group_by(excl_age, excl_miss_age, excl_miss_sts, excl_motivation) %>%
  summarize(n = n())
print(excl_all)

dat <-   alldat %>%
  filter(!id %in% c(excl_age$id, excl_miss_sts$id,
                    excl_miss_age$id, excl_motivation$id)) %>%
    mutate(sex = factor(sex, 1:2, c("m", "f")),
         ht = ht / 100,
         leg_m = leg / 100,
         leg_ht = leg_m / ht,
         bmi = wt / (ht * ht),
         agegrp = case_when(
           age %in% 5:7 ~ 1,
           age %in% 8:10 ~ 2,
           age %in% 11:13 ~ 3,
           age %in% 14:16 ~ 4),
         agegrp = factor(agegrp, 1:4, c("5-7", "8-10", "11-13", "14-16"))) %>%
```

```r
  mutate(sts2 = ifelse(sts2 > 90, NA, sts2),
         ht = ifelse(ht == 0, NA, ht),
         sts = map2_dbl(sts1, sts2, function(x, y) mean(c(x,y), na.rm = TRUE)),
         slj = pmap_dbl(list(slj1, slj2, slj3), function(x, y, z) mean(c(x,y,z), na.rm = TRUE)),
         watt = ifelse(maxtest == "yes", watt, NA)) %>%
  mutate(both_sts = !is.na(sts1) & !is.na(sts2),
         one_sts = xor(!is.na(sts1), !is.na(sts2)),
         any_shuttle = !is.na(shuttle),
         any_watt = !is.na(watt),
         any_slj = !is.na(slj)) %>%
  mutate(watt_kg = watt / wt,
         slj_ht = slj / ht,
         best_sts = map2_dbl(sts1, sts2, function(x, y) max(c(x,y), na.rm = TRUE)),
         sts_power = (leg / 100 - chair / 100) * wt * 9.81 * sts1 / 60,
         circumference = 2 * (chair / 100) * pi / 4,
         sts1_circ = sts1 * circumference)

zurich_id <- zurich %>%
  select(id) %>%
  mutate(Zurich = 1,
         id = as.character(id))
basel_id <- basel %>%
  select(id) %>%
  mutate(Basel = 1,
         id = as.character(id))
valid_id <- validdat %>%
  select(id) %>%
  mutate(Validity = 1,
         id = as.character(id))

wfawho2007<-read.table("wfawho2007.txt",header=T,sep="",skip=0) # only avail up to 10
hfawho2007<-read.table("hfawho2007.txt",header=T,sep="",skip=0)
bfawho2007<-read.table("bfawho2007.txt",header=T,sep="",skip=0)
source("who2007.r")

htdat <- dat %>%
  select(id, sex, age, ht, wt) %>%
  mutate(age_orig = age,
         age = age * 12 + 6,
         sex = unclass(sex),
         bmi = wt / (ht * ht),
         ht = ht * 100)
z1 <- left_join(htdat, hfawho2007, by = c("sex", "age")) %>%
  mutate(ht_z = (((ht / m)^l)-1) / (s * l)) %>%
  select(-l, -m, -s) %>%
  left_join(bfawho2007, by = c("sex", "age")) %>%
  mutate(bmi_z = (((bmi / m)^l)-1) / (s * l)) %>%
  select(-l, -m, -s, -ht, -wt, -bmi, -sex, -age)

dat <- full_join(dat, z1, by = "id")
med_iqr <- function(x, y, z, dig = 1){
  x <- format(x, digits = dig, nsmall = dig)
  y <- format(y, digits = dig, nsmall = dig)
```

```
  z <- format(z, digits = dig, nsmall = dig)
  paste(x, " [", y, ", ", z, "]", sep = "")
}

tab1 <- dat %>%
  group_by(agegrp) %>%
  summarize(n = n(),
            n_female = sum(sex == "f"),
            pct_female = mean(sex == "f"),
            wt_med = median(wt, na.rm = TRUE),
            wt_q1 = quantile(wt, 0.25, na.rm = TRUE),
            wt_q3 = quantile(wt, 0.75, na.rm = TRUE),
            ht_med = median(ht, na.rm = TRUE),
            ht_q1 = quantile(ht, 0.25, na.rm = TRUE),
            ht_q3 = quantile(ht, 0.75, na.rm = TRUE),
            htz_med = median(ht_z, na.rm = TRUE),
            htz_q1 = quantile(ht_z, 0.25, na.rm = TRUE),
            htz_q3 = quantile(ht_z, 0.75, na.rm = TRUE),
            bmi_med = median(bmi, na.rm = TRUE),
            bmi_q1 = quantile(bmi, 0.25, na.rm = TRUE),
            bmi_q3 = quantile(bmi, 0.75, na.rm = TRUE),
            bmiz_med = median(bmi_z, na.rm = TRUE),
            bmiz_q1 = quantile(bmi_z, 0.25, na.rm = TRUE),
            bmiz_q3 = quantile(bmi_z, 0.75, na.rm = TRUE),
            leg_med = median(leg_m, na.rm = TRUE),
            leg_q1 = quantile(leg_m, 0.25, na.rm = TRUE),
            leg_q3 = quantile(leg_m, 0.75, na.rm = TRUE),
            chair_med = median(chair, na.rm = TRUE),
            chair_q1 = quantile(chair, 0.25, na.rm = TRUE),
            chair_q3 = quantile(chair, 0.75, na.rm = TRUE)) %>%
  transmute(age = agegrp,
            n = as.character(n),
    sex_f = paste(n_female, " (", round(pct_female*100, 1), "%)", sep = ""),
    ht = pmap_chr(list(ht_med, ht_q1, ht_q3), med_iqr, dig = 2),
    ht_z = pmap_chr(list(htz_med, htz_q1, htz_q3), med_iqr, dig = 2),
    wt = pmap_chr(list(wt_med, wt_q1, wt_q3), med_iqr),
    leg = pmap_chr(list(leg_med, leg_q1, leg_q3), med_iqr, dig = 3),
    chair = pmap_chr(list(chair_med, chair_q1, chair_q3), med_iqr, dig = 1),
    BMI = pmap_chr(list(bmi_med, bmi_q1, bmi_q3), med_iqr),
    BMI_z = pmap_chr(list(bmiz_med, bmiz_q1, bmiz_q3), med_iqr))
tab1all <- dat %>%
  summarize(n = n(),
            n_female = sum(sex == "f"),
            pct_female = mean(sex == "f"),
            wt_med = median(wt, na.rm = TRUE),
            wt_q1 = quantile(wt, 0.25, na.rm = TRUE),
            wt_q3 = quantile(wt, 0.75, na.rm = TRUE),
            ht_med = median(ht, na.rm = TRUE),
            ht_q1 = quantile(ht, 0.25, na.rm = TRUE),
            ht_q3 = quantile(ht, 0.75, na.rm = TRUE),
            htz_med = median(ht_z, na.rm = TRUE),
            htz_q1 = quantile(ht_z, 0.25, na.rm = TRUE),
            htz_q3 = quantile(ht_z, 0.75, na.rm = TRUE),
```

```
            bmi_med = median(bmi, na.rm = TRUE),
            bmi_q1 = quantile(bmi, 0.25, na.rm = TRUE),
            bmi_q3 = quantile(bmi, 0.75, na.rm = TRUE),
            bmiz_med = median(bmi_z, na.rm = TRUE),
            bmiz_q1 = quantile(bmi_z, 0.25, na.rm = TRUE),
            bmiz_q3 = quantile(bmi_z, 0.75, na.rm = TRUE),
            leg_med = median(leg_m, na.rm = TRUE),
            leg_q1 = quantile(leg_m, 0.25, na.rm = TRUE),
            leg_q3 = quantile(leg_m, 0.75, na.rm = TRUE),
            chair_med = median(chair, na.rm = TRUE),
            chair_q1 = quantile(chair, 0.25, na.rm = TRUE),
            chair_q3 = quantile(chair, 0.75, na.rm = TRUE)) %>%
  transmute(age = "all",
            n = as.character(n),
    sex_f = paste(n_female, " (", round(pct_female*100, 1), "%)", sep = ""),
    ht = pmap_chr(list(ht_med, ht_q1, ht_q3), med_iqr, dig = 2),
    ht_z = pmap_chr(list(htz_med, htz_q1, htz_q3), med_iqr, dig = 2),
    wt = pmap_chr(list(wt_med, wt_q1, wt_q3), med_iqr),
    leg = pmap_chr(list(leg_med, leg_q1, leg_q3), med_iqr, dig = 3),
    chair = pmap_chr(list(chair_med, chair_q1, chair_q3), med_iqr, dig = 1),
    BMI = pmap_chr(list(bmi_med, bmi_q1, bmi_q3), med_iqr),
    BMI_z = pmap_chr(list(bmiz_med, bmiz_q1, bmiz_q3), med_iqr))
bind_rows(tab1, tab1all) %>%
  pivot_longer(cols = n:BMI_z) %>%
  pivot_wider(id_cols = name,
              names_from = age,
              values_from = value) %>%
  knitr::kable(caption = "Summary statistics for all data (including all participants 5-16 with at least 1

tab_S1 <- dat %>%
  group_by(agegrp, sex) %>%
  select(agegrp, sex, one_sts, both_sts, any_shuttle, any_watt, any_slj) %>%
  summarize(n = n(), across(everything(), ~ sum(.))) %>%
  mutate(any_sts = one_sts + both_sts) %>%
  select(agegrp, sex, n, any_sts, both_sts, one_sts, any_watt, any_slj)

summary(dat$chair)
tab2 <- dat %>%
  group_by(sex, agegrp) %>%
  summarize(n = n(),
            sts_p025 = quantile(best_sts, 0.025, na.rm = TRUE),
            sts_q1 = quantile(best_sts, 0.25, na.rm = TRUE),
            sts_med = quantile(best_sts, 0.5, na.rm = TRUE),
            sts_q3 = quantile(best_sts, 0.75, na.rm = TRUE),
            sts_p975 = quantile(best_sts, 0.975, na.rm = TRUE))
tab2all <- dat %>%
  group_by(sex) %>%
  summarize(n = n(),
            sts_p025 = quantile(best_sts, 0.025, na.rm = TRUE),
            sts_q1 = quantile(best_sts, 0.25, na.rm = TRUE),
            sts_med = quantile(best_sts, 0.5, na.rm = TRUE),
            sts_q3 = quantile(best_sts, 0.75, na.rm = TRUE),
            sts_p975 = quantile(best_sts, 0.975, na.rm = TRUE)) %>%
```

```r
  mutate(agegrp = "all")
tab2w <- bind_rows(tab2, tab2all) %>%
  rename(p025 = sts_p025,
         p25 = sts_q1,
         p50 = sts_med,
         p75 = sts_q3,
         p975 = sts_p975)
tab2f <- tab2w %>% filter(sex == "f") %>% ungroup() %>% select(-n, -sex) %>%
  rename_at(vars(p025:p975), ~ paste("F_", ., sep = ""))
tab2m <- tab2w %>% filter(sex == "m") %>% ungroup() %>% select(-n, -sex) %>%
  rename_at(vars(p025:p975), ~ paste("M_", ., sep = ""))

full_join(tab2f, tab2m) %>%
  knitr::kable(caption = "Reference values of the sit-to-stand test in children and adolescents (N = 547)"

dat %>%
  filter(age %in% 6:12) %>%
  group_by(sex) %>%
  summarize(n = n(),
            sts_mean = mean(best_sts, na.rm = TRUE),
            sts_p025 = quantile(best_sts, 0.025, na.rm = TRUE),
            sts_q1 = quantile(best_sts, 0.25, na.rm = TRUE),
            sts_med = quantile(best_sts, 0.5, na.rm = TRUE),
            sts_q3 = quantile(best_sts, 0.75, na.rm = TRUE),
            sts_p975 = quantile(best_sts, 0.975, na.rm = TRUE))
repsts <- dat %>%
  select(id, age, agegrp, sex, sts1, sts2) %>%
  mutate(mean_sts = (sts1 + sts2)/2,
         diff_sts = sts2 - sts1)
mean_diff <- mean(repsts$diff_sts, na.rm = TRUE)
sd_diff <- sd(repsts$diff_sts, na.rm = TRUE)
uloa <- mean_diff + 1.96 * sd_diff
lloa <- mean_diff - 1.96 * sd_diff
pcor <- function(x, y, z="1", meth = "pearson", plot = TRUE, fmt = FALSE, dig = 2,
                 xlbl = x, ylbl = y, zlbl = z, ...){
  cdat <- dat %>%
    select(any_of(c(x, y, z))) %>%
    drop_na()
  fm_x <- paste(x, "~", z)
  mod_x <- lm(as.formula(fm_x), data = cdat)
  fm_y <- paste(y, "~", z)
  mod_y <- lm(as.formula(fm_y), data = cdat)
  resx <- resid(mod_x)
  resy <- resid(mod_y)
  this_cor <- cor.test(resx, resy, method = meth, use = "pairwise")
  out <- format(this_cor$estimate, digits = dig, nsmall = dig)
  if(grepl("Pearson", this_cor$method)){
    out_ci <- format(c(this_cor$estimate, this_cor$conf.int), digits = dig, nsmall = dig)
    out <- paste(out_ci[1], " [", out_ci[2], " to ", out_ci[3], "]", sep = "")
  }
  if(plot){
    if(z != "1"){
    this_method <- ifelse(grepl("Pearson", this_cor$method), "Pearson", "Spearman")
```

```
    plot_title <- paste("r =", out)
    plot_subtitle <- paste("adjusted for", zlbl)
    plot_subtitle <- paste(this_method, ", ", plot_subtitle, sep = "")
    p <- qplot(resx, resy) +
      geom_smooth(method = "lm", formula = y ~ x, color = "black") +
      xlab(paste("residuals of", xlbl, "\nadjusting for", zlbl)) +
      ylab(paste("residuals of", ylbl, "\nadjusting for", zlbl)) +
      ggtitle(plot_title, subtitle = plot_subtitle) +
      theme(plot.title = element_text(size = rel(0.9)))
    return(p)
    } else {
    this_method <- ifelse(grepl("Pearson", this_cor$method), "Pearson", "Spearman")
    plot_title <- paste("r =", out)
    plot_subtitle <- "unadjusted"
    plot_subtitle <- paste(this_method, ", ", plot_subtitle, sep = "")
    p <- ggplot(aes(!!sym(x), !!sym(y)), data = cdat) +
      geom_point() +
      geom_smooth(method = "lm", formula = y ~ x, color = "black") +
      xlab(xlbl) + ylab(ylbl) +
      ggtitle(plot_title, subtitle = plot_subtitle) +
      theme(plot.title = element_text(size = rel(0.9)))
    return(p)
    }
  } else {
    if(grepl("Pearson", this_cor$method)){
    out <- c(this_cor$estimate, this_cor$conf.int)
    names(out) <- c("correlation", "lb", "ub")
    if(fmt){
      out_ci <- format(out, digits = dig, nsmall = dig)
      out <- paste(out_ci[1], " [95% CI ", out_ci[2], " to ", out_ci[3], "]", sep = "")
    }
    } else {
      out <- this_cor$estimate
      names(out) <- "correlation"
      if(fmt){
        out <- format(out, digits = dig, nsmall = dig)
      }
    }
    return(out)
  }
}


with(dat, cor(slj_ht, best_sts, use = "pairwise", method = "pearson"))
with(dat, cor(watt_kg, best_sts, use = "pairwise", method = "pearson"))

pcor("best_sts", "leg", "age", xlbl = "best STS", ylbl = "leg length")
pcor("best_sts", "leg", "1", xlbl = "best STS", ylbl = "leg length")
pcor("best_sts", "ht", "age", xlbl = "best STS", ylbl = "height")
pcor("best_sts", "ht", "1", xlbl = "best STS", ylbl = "height")
pcor("best_sts", "wt", "age", xlbl = "best STS", ylbl = "weight")
pcor("best_sts", "wt", "1", xlbl = "best STS", ylbl = "weight")
```

```r
pcor("best_sts", "watt_kg", xlbl = "best STS", ylbl = "Wmax / body weight")
pcor("best_sts", "slj_ht", xlbl = "best STS", ylbl = "SLJ / body height")


refadult <- tribble(
  ~sex,   ~agegrp, ~sts_p025, ~sts_q1, ~sts_med, ~sts_q3, ~sts_p975,
  "m", "20-24", 27, 41, 50, 57, 72,
  "m", "25-29", 29, 40, 48, 56, 74,
  "m", "30-34", 28, 40, 47, 56, 72,
  "m", "35-39", 27, 38, 47, 58, 72,
  "m", "40-44", 25, 37, 45, 53, 69,
  "m", "45-49", 25, 35, 44, 52, 70,
  "m", "50-54", 24, 35, 42, 53, 67,
  "m", "55-59", 22, 33, 41, 48, 63,
  "m", "60-64", 20, 31, 37, 46, 63,
  "m", "65-69", 20, 29, 35, 44, 60,
  "m", "70-74", 19, 27, 32, 40, 59,
  "m", "75-79", 16, 25, 30, 37, 56,
  "f", "20-24", 31, 39, 47, 55, 70,
  "f", "25-29", 30, 40, 47, 54, 68,
  "f", "30-34", 27, 37, 45, 51, 68,
  "f", "35-39", 25, 37, 42, 50, 63,
  "f", "40-44", 26, 35, 41, 48, 65,
  "f", "45-49", 25, 35, 41, 50, 63,
  "f", "50-54", 23, 33, 39, 47, 60,
  "f", "55-59", 21, 30, 36, 43, 61,
  "f", "60-64", 20, 28, 34, 40, 55,
  "f", "65-69", 19, 27, 33, 40, 53,
  "f", "70-74", 17, 25, 30, 36, 51,
  "f", "75-79", 13, 22, 27, 30, 43
  )  %>%
  mutate_at(vars(sex:agegrp), as_factor)

datref <- tab2 %>% select(-n) %>%
  bind_rows(refadult) %>%
  mutate(agegrp = factor(agegrp, c("5-7", "8-10", "11-13", "14-16", "17-19",
                                   paste(seq(20, 75, 5), seq(20, 75, 5) + 4, sep = "-")))) %>%
  ungroup() %>%
  mutate(sex = fct_recode(sex, "male" = "m", "female" = "f"))

datref <- datref %>%
  mutate(ageg = as.numeric(agegrp),
         stratum = as.numeric(agegrp %in% c("5-7", "8-10", "11-13", "14-16")),
         stratum = factor(stratum, 1:0, c("Children", "Adults")))
crs <- c("male" = "#dfc27d", "female" = "#80cdc1")
ggplot(aes(ageg, sts_med,
           ymin = sts_q1, ymax = sts_q3,
           color = sex, fill = sex, linetype = stratum), data = datref) +
  geom_line(size = 1.5) +
  geom_ribbon(alpha = 0.25) +
  geom_line(aes(ageg, sts_p025)) +
  geom_line(aes(ageg, sts_p975)) +
  annotate("segment", x = 1, xend = 4, y = 10, yend = 10) +
```

```
  annotate("text", x = 2.5, y = 14, label = "Reference Values\nin Children") +
  annotate("segment", x = 6, xend = 17, y = 10, yend = 10) +
  annotate("text", x = 11.5, y = 12, label = "Reference Values in Adults*") +
  xlab(NULL) +
  ylab("STS (repetitions)") +
  scale_x_continuous(breaks = 1:17, labels = levels(datref$agegrp)) +
  scale_linetype_manual(values = c("Children" = 1, "Adults" = 1)) +
  scale_color_manual(values = crs) +
  scale_fill_manual(values = crs) +
  guides(linetype = FALSE, color = guide_legend(NULL), fill = guide_legend(NULL)) +
  theme(legend.position = c(0.98, 0.98), legend.justification = c(1, 1))
p_sts12 <- ggplot(aes(sts1, sts2), data = repsts) +
  geom_abline(intercept = 0, slope = 1, color = "gray") +
  geom_point(size = 0.75) +
  scale_x_continuous(limits = c(25, 100)) + xlab("1st STS (repetitions)") +
  scale_y_continuous(limits = c(25, 100)) + ylab("2nd STS (repetitions)") +
  coord_equal() +
  labs(tag = "A")

p_ba <- ggplot(aes(mean_sts, diff_sts), data = repsts) +
  geom_point(size = 0.75) +
  geom_hline(yintercept = mean_diff) +
  geom_hline(yintercept = lloa, linetype = 2) +
  geom_hline(yintercept = uloa, linetype = 2) +
  xlab("Mean of STS Measurements") + ylab("Difference in STS measurements") +
  annotate("text", x = 31, y = mean_diff + 2,
           label = paste("", round(mean_diff, 1))) +
  annotate("text", x = 31, y = uloa + 2,
           label = paste("",
                         round(uloa, 1))) +
  annotate("text", x = 31, y = lloa + 2,
           label = paste("",
                         round(lloa, 1))) +
  labs(tag = "B")

p_sts12 + p_ba
pcor("watt_kg", "best_sts", "1",
     xlbl = "Maximal exercise test (Wmax / body weight)", ylbl = "Best STS") +
    labs(tag = "A") +
pcor("slj_ht", "best_sts", "1",
     xlbl = "SLJ (distance / body height)", ylbl = "Best STS") +
  labs(tag = "B")

tab1b <- dat %>%
  group_by(sex, agegrp) %>%
  summarize(n = n(),
            p_STS_2 = sum(both_sts),
            p_SLJ = sum(any_slj),
            p_Wmax = sum(any_watt))
tab1b_all <- dat %>%
  summarize(n = n(),
            p_STS_2 = sum(both_sts),
            p_SLJ = sum(any_slj),
```

```
            p_Wmax = sum(any_watt))%>%
  mutate(agegrp = "all", sex = "all")

bind_rows(tab1b, tab1b_all) %>%
  mutate_at(vars(p_STS_2:p_Wmax), ~ paste(round(. * 100 / n), "%", sep = "")) %>%
  knitr::kable(caption = "Participation in other measures of physical fitness by age and sex")
dat_sts <- dat %>%
  select(id, agegrp, age, sex, sts1, sts2) %>%
  mutate(mean_sts = map2_dbl(sts1, sts2, function(x, y) mean(c(x,y), na.rm = TRUE)),
        best_sts = map2_dbl(sts1, sts2, function(x, y) max(c(x,y), na.rm = TRUE)),
        first_sts = sts1,
        last_sts = map2_dbl(sts1, sts2, function(x, y) ifelse(!is.na(y), y, x))) %>%
  pivot_longer(mean_sts:last_sts,
               names_to = "definition",
               names_pattern = "(.*)_sts",
               values_to = "sts") %>%
  group_by(definition) %>%
  mutate(mean = mean(sts))

myf <- function(x){format(x, digits = 0, nsmall = 0)}
summ_sts <- dat_sts %>%
  group_by(sex, agegrp, definition) %>%
  summarize(median = median(sts),
            q1 = quantile(sts, p = 0.25),
            q3 = quantile(sts, p = 0.75),
            p025 = quantile(sts, p = 0.025),
            p975 = quantile(sts, p = 0.975),
            summary = paste(myf(median), " [", myf(q1), " to ", myf(q3), "]", sep = "")) %>%
  pivot_wider(id_cols = sex:agegrp, names_from = definition, values_from = summary)

summ_sts %>%
  select(sex, agegrp, first, best, last, mean) %>%
  knitr::kable(caption = "Reference values (median [Q1 to Q3], by sex, age group and definition of STS")

tmp <- dat %>%
  select(id, agegrp, age, sex, sts1, sts2, both_sts) %>%
  mutate(mean_sts = map2_dbl(sts1, sts2, function(x, y) mean(c(x,y), na.rm = TRUE)),
        best_sts = map2_dbl(sts1, sts2, function(x, y) max(c(x,y), na.rm = TRUE)),
        first_sts = sts1,
        last_sts = map2_dbl(sts1, sts2, function(x, y) ifelse(!is.na(y), y, x))) %>%
    pivot_longer(mean_sts:last_sts,
               names_to = "definition",
               names_pattern = "(.*)_sts",
               values_to = "sts") %>%
  group_by(sex, agegrp, definition) %>%
  summarize(p50_all = quantile(sts, p = 0.5),
            q1_all = quantile(sts, p = 0.25),
            q3_all = quantile(sts, p = 0.75),
            p50_both = quantile(sts[both_sts], p = 0.5),
            q1_both = quantile(sts[both_sts], p = 0.25),
            q3_both = quantile(sts[both_sts], p = 0.75)) %>%
  pivot_longer(p50_all:q3_both,
               names_to = c("qu", "subset"),
```

```
                names_sep = "_") %>%
  pivot_wider(names_from = qu,
              values_from = value)

tmp %>% filter(definition == "best")

# ggplot(aes(agegrp, p50, ymin = q1, ymax = q3,
#             color = definition, linetype = subset),
#         data = tmp) +
#   geom_linerange(position = position_dodge(width = 0.75)) +
#   geom_point(position = position_dodge(width = 0.75)) +
#   facet_wrap(vars(sex)) +
#   xlab(NULL) + ylab("STS (median, Q1 to Q3)") +
#   scale_color_brewer(palette = "Dark2") +
#   scale_y_continuous(breaks = seq(40, 80, 10),
#                      minor_breaks = seq(40, 80, 2))
age_scale <- scale_x_continuous(breaks = seq(6, 15, 3),
                                minor_breaks = seq(5, 16, 1))

p_age_sts <- pcor("age", "best_sts", ylbl = "Best STS") + age_scale + labs(tag = "A")
p_age_slj <- pcor("age", "slj_ht", ylbl = "SLJ (distance / body height)") + age_scale + labs(tag = "B")
p_age_watt <- pcor("age", "watt_kg", ylbl = "Maximal exercise test (Wmax / body weight)") + age_scale + la

p_age_sts + p_age_slj + p_age_watt
p1 <- pcor("watt_kg", "sts1", "age",
           xlbl = "Maximal exercise test\n(watt / weight)", ylbl = "STS") +
  labs(tag = "A")
p2 <- pcor("slj_ht", "sts1", "age",
           xlbl = "SLJ (distance / height)", ylbl = "STS") +
  labs(tag = "B")
p1 +  p2
p1 <- pcor("ht", "best_sts",  xlbl = "height", ylbl = "Best STS")
p2 <- pcor("leg", "best_sts",  xlbl = "leg length", ylbl = "Best STS")
p3 <- pcor("leg_ht", "best_sts",  xlbl = "leg length / height", ylbl = "Best STS")
p1b <- pcor("ht", "best_sts", "age", xlbl = "height", ylbl = "Best STS")
p2b <- pcor("leg", "best_sts", "age", xlbl = "leg length", ylbl = "Best STS")
p3b <- pcor("leg_ht", "best_sts", "age", xlbl = "leg length / height", ylbl = "Best STS")
p1 + p2 + p3 +
  p1b + p2b + p3b
si <- sessionInfo()
rver <- si$R.version$version.string
pk1 <- si$basePkgs
pk2 <- si$otherPkgs
pk2v <- NULL
for (package_name in names(pk2)){
     pk2v <- c(pk2v, paste(package_name, packageVersion(package_name)))
}
```

#### Computational Details

- R version: R version 4.0.3 (2020-10-10)
- Base packages: stats, graphics, grDevices, utils, datasets, methods, base
- Other packages: patchwork 1.0.1, tableone 0.12.0, broom 0.7.5, readxl 1.3.1, forcats 0.5.0, stringr 1.4.0, dplyr

1.0.5, purrr 0.3.4, readr 1.4.0, tidyr 1.1.3, tibble 3.1.0, ggplot2 3.3.3, tidyverse 1.3.0

This document was generated on 2021-03-23 at 13:43.
